# Predicting Active Suicidality from Temporal Dynamics of Implicit Life-Death Associations

**DOI:** 10.64898/2026.09.09.26362642

**Authors:** Pedram Rajaei, Aaron N. McInnes, Benito M. Garcia, Shreya Yadav, Blair Brown, Miriam Freedman, Sophia M Balint, Melanie D. Goodman Keiser, Shahabedin Tavasoli, Michael V. Bronstein, Alik S. Widge, Ali Yousefi

**Affiliations:** Department of Biomedical Engineering, University of Houston, Houston, TX, USA; Department of Psychiatry and Behavioral Sciences, University of Minnesota, Minneapolis, MN, USA; Institute for Health Informatics, University of Minnesota, Minneapolis, MN, USA

**Keywords:** Suicidal ideation, Implicit association test, Reaction time, Ecological momentary assessment, Machine learning, Cognitive control

## Abstract

**Background:** Interview-based suicide assessments are insufficient for near-term risk detection, but there are no evidence-based, high-performing alternatives. The Death Implicit Association Test (DIAT) has been proposed as a suicide risk assessment tool. However, traditional DIAT scoring methods exhibit limited association with suicidality.

**Methods:** We tested whether temporal structure in reaction times (RTs) from the Brief Death Implicit Association Test (BDIAT) tracks active suicidal ideation (SI), which was ascertained by ecological momentary assessment (EMA). Seventy-seven participants who either did or did not endorse active SI during EMA (SI+ n=22; SI− n=55) completed 18 alternating blocks of the BDIAT (9 Life + Me, 9 Death + Me). We computed the conventional D-score, an aggregate index of self-life versus self-death associations, and separately treated RTs as time-varying signals, examining entrainment to block alternation and habituation during conflict-related trials. We then developed a bilinear logistic regression classifier to extract these temporal RT features and trained it to predict SI status, with performance validated using leave-one-out cross-validation.

**Results:** The conventional D-score did not reliably classify SI+/SI− individuals. In contrast, temporal RT dynamics reveal group differences in block-level entrainment and trial-level habituation. A bilinear logistic regression classifier leveraging these dynamics predicts SI status with 77% balanced accuracy and AUC 0.799.

**Conclusions:** Results demonstrate that temporal dynamics of RT during the BDIAT track active ideation and can be used to distinguish SI status at the individual level, implicating RT temporal dynamics evoked during BDIAT as a candidate behavioral marker of near-term suicide risk.

## Introduction

Clinicians consider active suicidal ideation (SI), thoughts of ending one’s life that include method, plan, and/or intent(1), a particularly important marker of suicide risk(2). Identifying people with active SI is therefore essential for risk mitigation, but remains difficult in clinical practice. Clinicians lack objective, behavior-based markers that can track near-term risk in real time. They therefore often rely on self-report assessments, such as the Columbia Suicide Severity Rating Scale (C-SSRS), that are subject to recall bias and vulnerable to purposeful non-disclosure, which is common due to factors including concern about loss of autonomy(2,3). These assessments also cannot scale to meet demand. Thoughts of suicide are extremely common, impacting over 10 million adults per year in the United States alone(4). Screening these individuals with the few existing well-trained C-SSRS interviewers is not feasible, particularly because SI can rapidly fluctuate across short timescales (hours, days)(2,3,5–7), requiring repeated assessment to reveal when people are at risk(8). Assessment methods that are more scalable, robust to purposeful non-disclosure, and compatible with within-person variation in ideation could save lives by enabling timely interventions(5).

The Brief Death Implicit Association Test (BDIAT; **Figs. 1A-B**) has emerged as a potential platform for objective, computerized, and behavior-based suicide assessment(9). Traditionally, reaction times (RTs) on this task are used to produce a difference score, also called a “D-score,” which indexes the strength of implicit death-self associations(9). This D-score shows modest but meaningful ability to distinguish individuals with and without various histories of suicide-related thoughts and behaviors, including people who differ on levels of active suicidal ideation(10,11). Nevertheless, the D-score’s utility for identifying near-term suicide risk or informing clinical decision-making remains limited and varies substantially across settings(10,11). The poor performance of D-scores is possibly due in part to their reliance on RT averages and the resulting loss of information in trial-by-trial variability in RTs(12,13), even though (1) such variability and its temporal dynamics can carry meaningful information about underlying cognitive processes in psychophysical tasks(14,15) and (2) there is strong reason to expect that trial-by-trial variability in BDIAT RTs will differ systematically according to one’s level of suicidality, providing a marker of near-term ideation and suicide risk.

**Figure 1.**
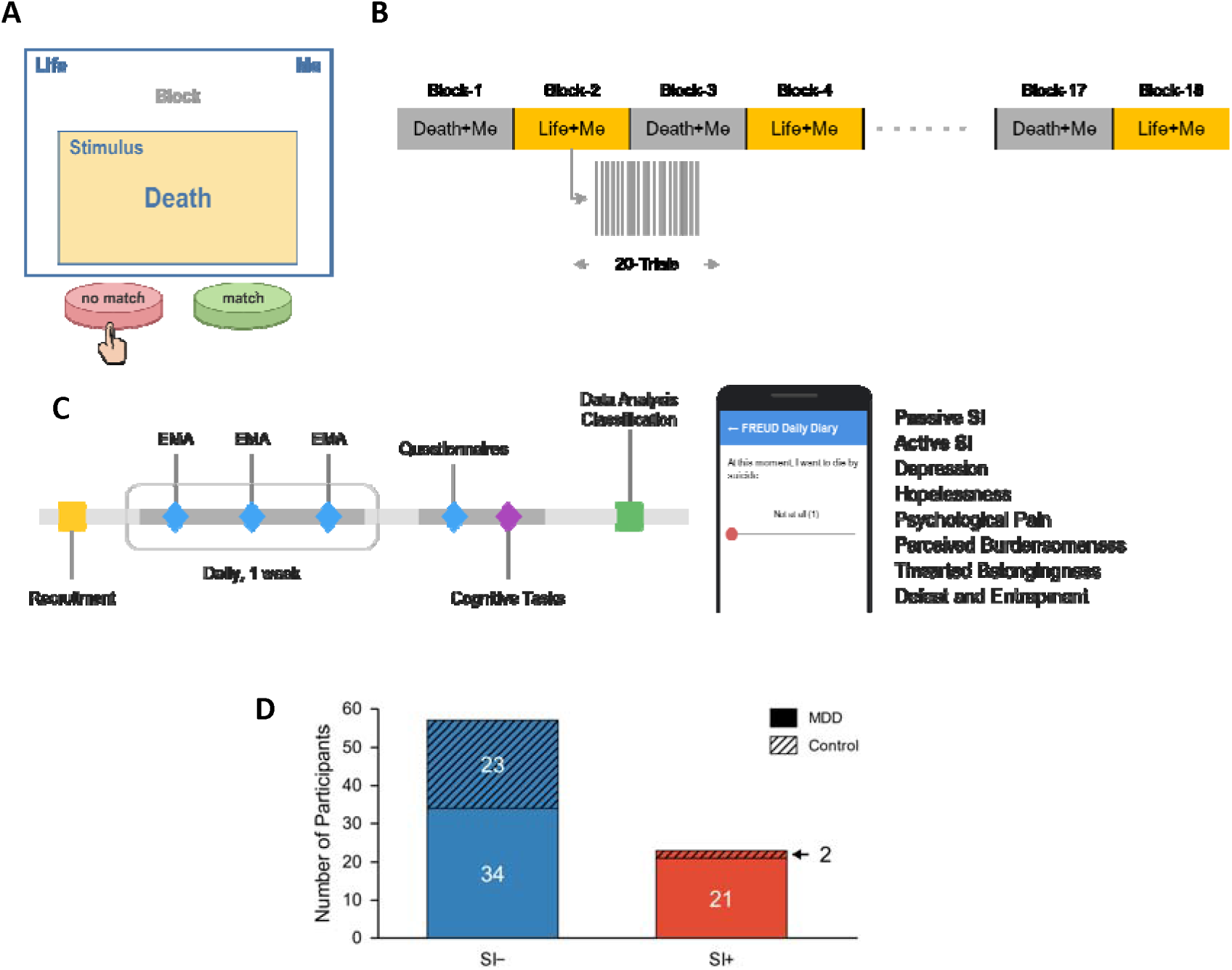
Brief Death Implicit Association Task (BDIAT) and Ecological Momentary Assessment (EMA). **(A)** Example trial from the BDIAT. The category labels (“Life” and “Me”) appear at the top of the screen, while the stimulus word changes on each trial. In this example, the stimulus is the word “Death.” Participants indicate whether the stimulus is semantically associated with either of the category labels by responding “match” or “no match” using designated keyboard keys. In this case, the correct response is “no match.” Reaction time (RT) and response accuracy are recorded on each trial. Participants are given up to 2.0 seconds to respond before the task advances to the next trial. Stimulus words, such as “I,” “They,” “Death,” and “Life,” are presented in random order across trials. **(B)** The BDIAT consists of 360 trials divided into 18 blocks of 20 trials, alternating between “Death + Me” and “Life + Me” blocks. Each trial lasts up to 2.0 seconds, with an inter-trial interval of approximately 500 milliseconds, yielding a total task duration of approximately 14 minutes. **(C) Left**: Timeline of the longitudinal study, showing the temporal alignment of daily behavioral task sessions and EMA administrations. **Right**: EMAs were administered three times daily to assess momentary fluctuations in suicidal ideation and related psychological symptoms, including psychological pain. This sampling approach enabled the mapping of discrete task performance onto high-frequency clinical measures. **(D)** Distribution of participants by diagnostic group and suicidal ideation status. Bars show the numbers of participants classified as SI+ (with active suicidal ideation) or SI− (without active suicidal ideation), stratified by major depressive disorder (MDD) and control diagnosis. Two participants in the SI− group and one participant in the SI+ group were excluded from the analysis because of missing or censored data.

This variability may reflect differences in conflict monitoring across people with and without active SI. According to conflict monitoring theory, when a stimulus elicits competing, incompatible responses, the brain detects this conflict and recruits cognitive control, producing a transient slowing of RT known as conflict adaptation before performance returns to baseline(16). It follows from this theory that aberrant patterns of RT slowing may result from the deficits in conflict processing and related neurocircuitry observed in people with histories of suicidal thoughts and behaviors(17,18). Different slowing patterns in people with/without active ideation may also stem from variation in the amount of conflict that BDIAT stimuli elicit. A “self-life” pairing in the task may elicit conflict for an individual with active SI, whereas a “self-death” pairing may do the same for someone without active SI. That is, conflict adaptation may be triggered on certain BDIAT trials only for people with particular histories of suicidality. If the timing of adaptive slowing within BDIAT blocks, and its evolution across multiple trials, does indeed differ systematically between people with and without active SI, one might expect SI-related differences in BDIAT performance to appear most clearly during transitions across trial and block types, rather than in aggregate RT measures, such as D-scores.

Trial-to-trial variability in BDIAT has not been extensively explored, largely for methodological reasons. Many methods that could measure trial-by-trial RT variability to identify people with SI are poorly suited to the small, noisy datasets typically available from BDIAT studies in clinical populations. Sequence-based deep learning approaches, including sequence-to-sequence models(19) and transformer-based architectures(20), are generally designed for larger datasets than those available in this setting. Moreover, linking these methods’ predictions to interpretable behavioral and physiological mechanisms, an essential requirement for clinical decision-making(21), is not straightforward and often requires extensive post hoc analysis. Even when these approaches are used as embedding pipelines to extract behavioral features for downstream prediction, their utility remains limited in small-sample settings. Alternative solutions are therefore needed to extract temporally structured information from IAT performance in a manner that is both clinically meaningful and compatible with limited data availability.

With this in mind, we paired ecological momentary assessment (EMA) of near-term active SI with trial- and block-level analyses of RTs recorded during a modified BDIAT. We tested whether conflict adaptation and implicit self-referential processing exhibit distinct temporal signatures in individuals with active SI (SI+; n=22) compared with those without active SI (SI−; n=55). While we had strong reason to expect that such signatures would not be captured by average RTs (or D-scores) alone, previously published research provided fewer hints about the optimal time window across which variability would differentiate people with varying levels of ideation. Accordingly, we examined temporal structure across multiple scales, including block-level entrainment to the alternating task design, trial-level adaptation following block switches, and compact latent representations of RT dynamics. We then used logistic regression to evaluate whether these predefined temporal features could distinguish SI+ from SI− participants while allowing each feature’s contribution to the prediction to be examined directly. This approach treats BDIAT performance not as a single implicit-association score, but as a time-varying behavioral signal whose dynamics may reveal clinically relevant cognitive processes associated with near-term ideation and suicide risk.

## Methods

### Participants

Participants (age 18–65) were recruited as part of a larger study on neurobehavioral markers of suicide risk; the cohort includes individuals with treatment-resistant depression recruited from interventional psychiatry clinics and a psychiatrically healthy comparison group. All participants provided written informed consent under protocols approved by the Advarra IRB and U.S. Navy HRPO. Full eligibility and recruitment criteria are provided in Supplementary Information, **Section 1**; enrollment numbers and data-quality exclusions are detailed in **Section 2**.

### EMA and clinical measures

Participants completed EMA surveys measuring active and passive suicidal ideation three times daily for seven days prior to the BDIAT session. Participants were classified as SI+ if their mean active-SI score exceeded zero, and SI− otherwise. This EMA-derived binary label served as the primary outcome for all analyses; this assumption is consistent with growing evidence supporting EMA as a sensitive method for capturing near-term suicidal ideation(22,23). The C-SSRS(2) was administered as a secondary clinical benchmark. Several participants who did not endorse active ideation on the C-SSRS reported active suicidal thoughts on their EMAs. This pattern is consistent with the notion that single-timepoint assessments miss transient ideation that repeated sampling captures. Full details of participant classification counts, and the C-SSRS/EMA comparison are provided in Supplementary Information, **Section 3**.

### Behavioral task

The BDIAT, which was administered approximately one week after the EMA period, consisted of 360 trials across 18 alternating “Life + Me” and “Death + Me” blocks (20 trials/block; Supplementary Information, **Section 4**). An independent online validation cohort completed a variant with 36 blocks of 10 trials each (Supplementary Information, **Section 5**).

### Analysis overview

We first computed the conventional D-score, a standardized RT difference between block types (Supplementary Information, **Section 6**). RT sequences were then pre-processed using a state-space filtering approach(24,25), which gives a denoised (smooth), baseline-normalized RT signal. Missing or censored trials were handled using different imputation strategies; we confirmed our results were robust to this choice (Supplementary Information, **Sections 7–8**). We also looked at block-level rhythmicity using an autocorrelation-based Rhythm Index and checked the significance of this rhythmic pattern by comparing it against shuffled versions of the data (Supplementary Information, **Sections 9–10**). At the trial level, we used mixed-effects modeling (Supplementary Information, **Section 12**) and a PCA-based approach applied to “Death + Me” block trials to capture how RTs evolved within a block (Supplementary Information, **Section 13**). We also checked that these PCA results held up when RTs were represented on different scales: raw, log-transformed, and inverse-power (Supplementary Information, **Section 14**). Separately, we ruled out the possibility that temporal dynamics in reaction times could be explained by the frequency with which stimulus words happened to appear early versus late in the task (Supplementary Information, **Section 15**). Finally, we built a classifier that combines both the block-level and trial-level RT patterns to predict SI status (SI+ vs. SI−), and tested how it performed using leave-one-out cross-validation and permutation tests. Additional comparisons with other classifiers and checks designed to provide insight into the specific features of reaction time patterns that were most important to model decisions are provided in Supplementary Information, **Sections 16–19**.

## Results

### D-Scores Yield No Distinction Between SI+ and SI− Participants

For each participant, we computed the conventional BDIAT D-score(9), defined as the standardized difference in mean RTs between “Death + Me” and “Life + Me” blocks, in keeping with established IAT procedures(12,26). Details of D-score computation are provided in the Supplementary Information, **Section 6**. The final analytic sample included 55 SI− and 22 SI+ participants; participant characteristics, data collection, and preprocessing procedures are also described in the Supplementary Information, **Section 7**. In our sample, D-scores did not differ significantly between SI+ and SI− participants. Using the D-score alone to classify SI status (SI+ versus SI−, with SI+ treated as the positive class) yielded near-chance performance, with balanced accuracy = 45.9%, sensitivity = 40.9%, specificity = 50.9%, and area under curve (AUC) = 0.505 (**Figs. 2A-B**). This contrasts with results from other studies, which reported modest discrimination suicide-risk groups, with AUC values typically ranging from 0.60 to 0.64 (Cohen’s d ≈ 0.30–0.45)(27,28) (for potential explanations, see Discussion).

**Figure 2.**
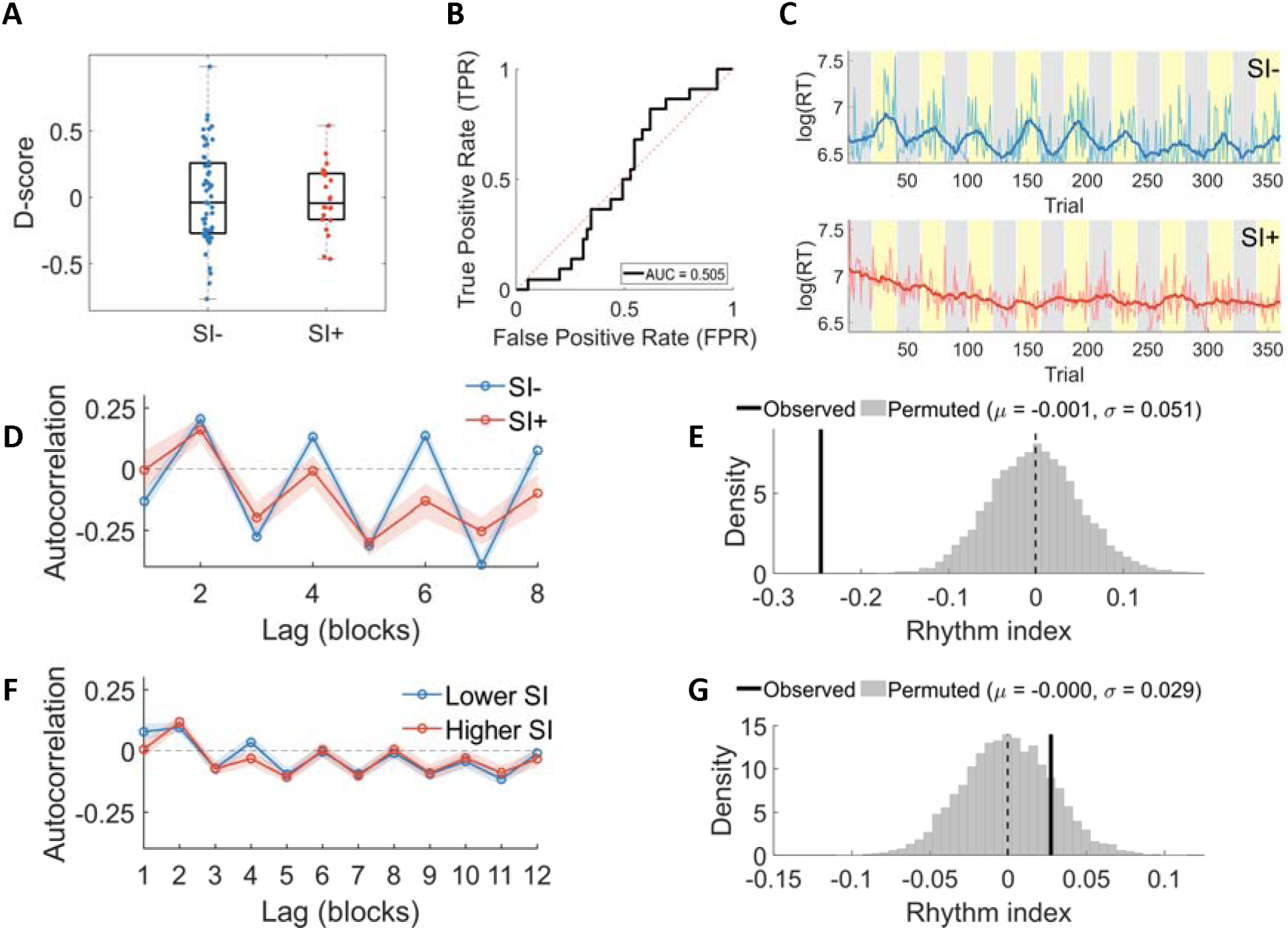
D-score Prediction Performance and Block-level Temporal Structure. **(A)** Box plot of D-scores for participants with active suicidal ideation (SI+) and without active suicidal ideation (SI−) in the primary cohort (n = 77); substantial overlap indicates minimal discriminative power. **(B)** Receiver operating characteristic (ROC) curve for SI classification based on the D-score. Participants were classified as SI+ when D > 0. The area under the curve (AUC = 0.505) indicates near-chance predictive performance. Under this decision rule, sensitivity was 40.9% and specificity was 50.9%. **(C)** Example RT profiles from one SI-participant (top) and one SI+ participant (bottom). Log-transformed RTs are shown in light cyan, and the filtered RT trace estimated by the state-space model (see Supplementary Information, **Section 7**) is shown in blue. Gray and yellow stripes denote “Death + Me” and “Life + Me” blocks, respectively. The SI-participant shows clear modulation of RT as the task switches between block types, whereas this pattern is less apparent in the SI+ participant, potentially suggesting altered conflict adaptation. **(D)** Block-level autocorrelation function (ACF; mean ± 95% bootstrap confidence intervals) across eight lags, computed from mean reaction time (RT) per block in the primary cohort. RTs were mean-centered and linearly detrended within participant before autocorrelation was computed. SI− participants show a rhythmic pattern, with positive correlations at even lags and negative at odd, consistent with entrainment to the alternating block structure. In contrast, SI+ participants exhibit weaker and shorter-lived autocorrelation. **(E)** Null distribution of the group difference in the Rhythm Index (see main text) obtained after shuffling block order in the primary cohort (n = 77). The observed difference from the true block order (black line) lies outside the null distribution centered near zero, indicating that the rhythmic structure in **D** depends on the task-defined block arrangement rather than nonspecific temporal dependencies. **(F)** Block-level ACF (mean ± 95% bootstrap confidence intervals) from an independent online implementation of the BDIAT using shorter blocks (10 trials per block; 36 alternating “Death + Me” and “Life + Me” blocks) (n = 83). Participants were grouped into lower and higher suicidal-ideation groups based on self-reported C-SSRS responses, approximating the SI− and SI+ grouping used in the primary cohort. RTs were detrended and mean-centered within participant before analysis. Block-level ACFs show a similar oscillatory structure to the primary cohort, although correlations are weaker. **(G)** Null distribution of the group difference in the Rhythm Index after shuffling block order in the online cohort (n= 83). The observed group difference from the true data (black line) falls within the null distribution, consistent with the absence of a significant group effect (p = 0.34). Together with the nonsignificant between-group comparison (p = 0.56), this suggests that although rhythmic structure is present at the subject level, it does not reliably distinguish lower versus higher suicidal-ideation groups in the online sample.

### Rhythmic Entrainment to Block Structure Is Present in BDIAT and Differs Across SI Groups

The BDIAT alternates between “Death + Me” and “Life + Me” blocks. The logic underlying the D-score dictates that differences in RTs across these block types carry information about suicidality and other outcomes. Analyses focused on the alignment of RTs to block type alternation might extract this information without losing information to whole-experiment averaging as D-scores do. As a first step, we confirmed that participants’ RTs showed block-locked modulation across the alternating task conditions (example: **Fig. 2C**). Building on our preliminary spectral findings from a smaller sample(29), we used block-level autocorrelation to quantify task-locked RT dynamics in the larger EMA-characterized cohort. For each participant, we averaged baseline-scaled RTs (RT-scale) across trials within each block to obtain a block-level time series of mean RT-scale values. Block-level autocorrelation functions (ACFs) revealed a clear oscillatory pattern consistent with the alternating block design, characterized by negative autocorrelation at odd lags and positive autocorrelation at even lags (**Fig. 2D**). To quantify this effect, we computed a Rhythm Index, defined as the mean difference between even-lag and odd-lag autocorrelations (Supplementary Information, **Section 9**). This scalar index summarizes ABAB entrainment, with larger values indicating stronger alignment of RT dynamics with the alternating task structure. By averaging this contrast across multiple lags, the Rhythm Index reduces sensitivity to idiosyncratic fluctuations at any single block transition or autocorrelation lag.

To rigorously test whether there was an oscillating pattern to participant RTs across blocks, we compared the Rhythm Index calculated from our data to that obtained under a within-subject shuffle that destroyed the ABAB block order. The Rhythm Index was significantly greater in our real data (p = 5.87 × 10⁻¹²; Supplementary Information, **Fig. S3A**), demonstrating task-locked rhythmicity. The Rhythm Index was also greater under shuffles that preserved ABAB structure while randomizing block order within category (p = 0.0068; Supplementary Information, **Fig. S3B**), indicating that the effect was not explained by slow drift, practice effects, or block identity alone.

Next, we examined whether the Rhythm Index carried information about suicidality. SI− participants exhibited a larger Rhythm Index than SI+ participants (Welch’s t-test, t(41.4) = −2.30, p = 0.0269), indicating stronger behavioral alignment to task structure. The observed SI− versus SI+ difference also lay outside the null distribution generated by the destroy-ABAB shuffle (p < 0.001; **Fig. 2E**), supporting the conclusion that this group difference in task-locked block-level RT dynamics is meaningful and not due to chance. This result supports the intuition underlying the D-score – that RT differences across blocks carry information about suicidality.

Given this finding, we wondered whether our particular choices of task setting (in person vs. online) and BDIAT block number and length were optimal for distinguishing SI+ and SI− via the Rhythm Index. As an initial step toward answering this question, we examined whether RT rhythms were block-locked and SI-associated in other BDIAT instantiations. If so, these could be searched systematically to reveal which yielded the best discrimination. To begin, we administered an online version of the BDIAT using shorter 10-trial blocks in an independent cohort. Details of the online validation cohort are provided in the Supplementary Information, **Section 5**. Eighty-three participants were classified into lower- and higher-SI groups using self-reported C-SSRS responses(2). Autocorrelation analyses revealed a block-locked rhythmic structure consistent with the primary cohort (**Fig. 2F**). Rhythm Index values again exceeded both destroy-ABAB and preserve-ABAB shuffle nulls (Supplementary Information, **Figs. S3C-D**). However, group-label permutation tests did not show a reliable group difference in Rhythm Index (p = 0.56), nor did destroy-ABAB block permutations (p = 0.34; **Fig. 2G**). These findings indicate that entrainment to block-level task context generalizes across task parameters and data-collection settings, whereas SI-related group discrimination was evident only in the EMA-characterized cohort. Ideally, our approach to discriminating more and less suicidal people should work regardless of differences in administration mode or (within reason) task parameters – enabling assessment to be convenient and robust to common occurrences in applied settings (e.g., a patient stops the task early to meet with a clinician).

We posited that block-level rhythms might be limited in their discrimination of suicidality levels because aggregate metrics obscure the transient cognitive conflicts associated with active SI. We reasoned that a focus on moment-to-moment transitions might better isolate the impacts of these transient conflicts on RTs, exposing more robust group differences. Accordingly, we next examined fine-grained, trial-level RT dynamics to capture the immediate behavioral adjustments following block switches.

### Temporal Encoding of SI State Is Evident in Trial-level RT Dynamics

To characterize trial-wise adjustments, we used RT-scale, which quantifies proportional deviations from each participant’s estimated response-speed baseline after preprocessing missing and censored RTs. Details of preprocessing and normalization are provided in the Supplementary Information, **Section 7**.

**Figs. 3A** and **3B** show the group-level mean RT-scale trajectories in “Death + Me” and “Life + Me” blocks across the 20 trials of each block. On the first trial, both groups showed a marked RT elevation, with mean RT-scale values of approximately 1.25, corresponding to a ~25% increase over baseline. In “Death + Me” blocks, mean RT-scale values returned toward baseline by trials 2–3. Beginning at trial 4, however, the groups’ trajectories began to diverge, with SI− participants showing lower RT-scale values across trials 4–6. A similar but weaker pattern was observed in “Life + Me” blocks. These trial-wise differences were suggestive but did not survive Benjamini–Hochberg false discovery rate (FDR) correction at individual trials (all q-values > 0.05)(30).

**Figure 3.**
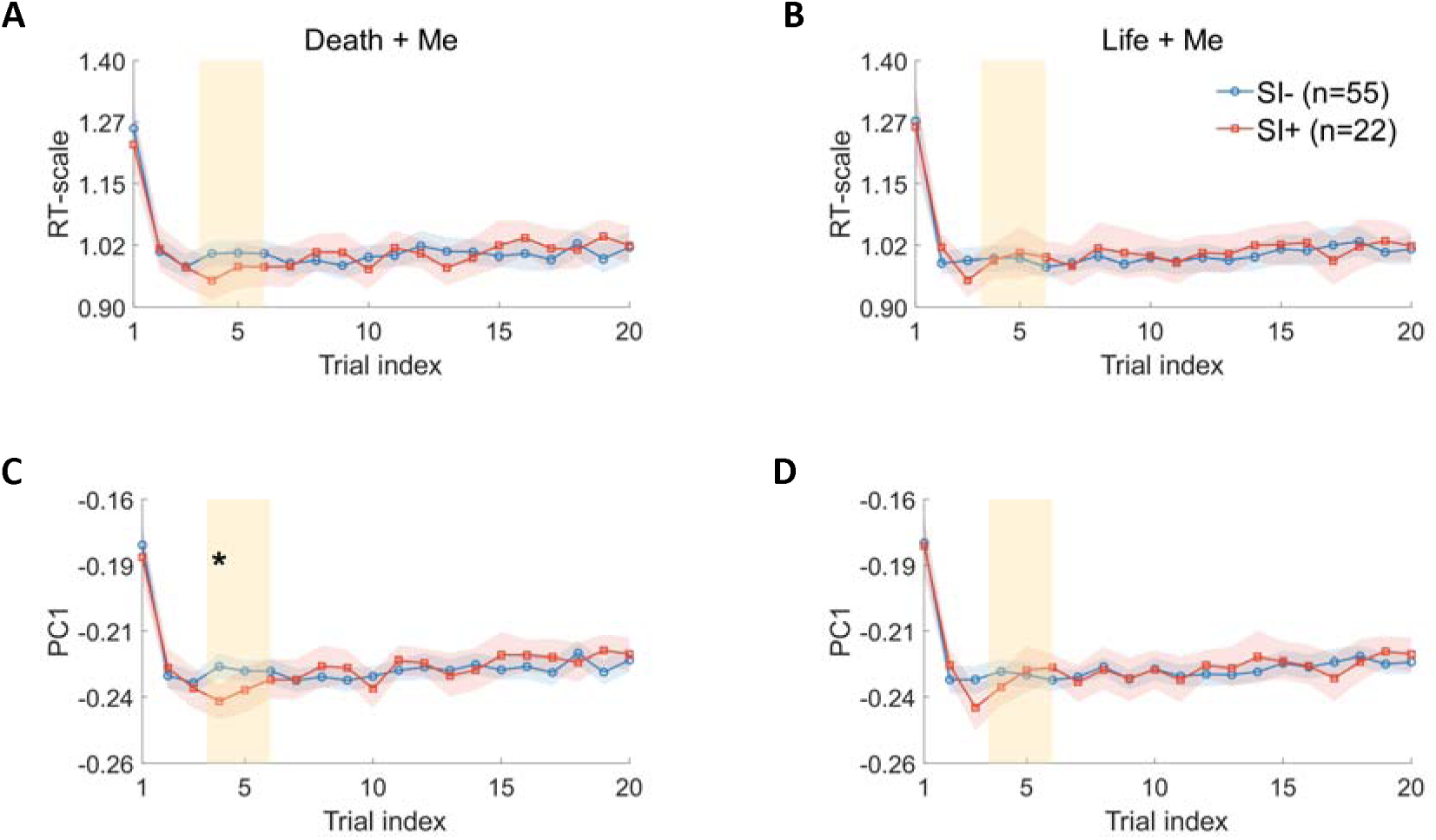
Evidence for Temporal Encoding of Suicidal Ideation (SI) State in Reaction Time (RT). **(A)** Mean baseline-scaled RT (RT-scale) across trials in “Death + Me” blocks, shown separately for participants with active suicidal ideation (SI+, red line with squares) and without active suicidal ideation (SI−, blue line with circles); shaded bands represent bootstrap 95% confidence intervals. Both groups show a pronounced elevation on the first trial followed by rapid return toward baseline, consistent with an initial switch cost. Although the trajectories show modest descriptive divergence at several trial positions, none of the trial-wise group differences are significant after correction for multiple comparisons. **(B)** Mean RT-scale across trials in the “Life + Me” blocks. As in **A**, both groups show rapid normalization after the first trial, with little evidence for sustained group separation across the remainder of the block. **(C)** Group-averaged PC1 trajectory derived from trial-wise RT-scale patterns in the “Death + Me” blocks using uncentered principal component analysis (PCA). The largest group separation emerges around trial 4, where the between-group difference survives FDR correction (*q* ≤ 0.05). **(D)** Same analysis as in **C**, but for the “Life + Me” blocks. In contrast to the “Death + Me” condition, the PC1 trajectory shows weaker and less temporally localized group differentiation. Yellow shading in all panels marks the early-trial window (trials 4–6), and the asterisk indicates the trial with an FDR-corrected group difference. Higher RT-scale values indicate slower responses relative to each participant’s baseline RT. Trial-wise statistics for all panels are provided in Supplementary Information, **Fig. S8.**

A test of the overall temporal structure of RT adjustments across the block might be better powered (due to eliminating the need for mass FDR correction) and better able to discern group differences that play out across multiple trials. We therefore tested whether SI state influenced the overall temporal structure of RT adjustments across the block. Linear mixed-effects models were fit separately for “Death + Me” and “Life + Me” blocks, with Group, Trial, and their interaction as fixed effects and participant-level random intercepts. In “Death + Me” blocks, this analysis revealed a significant Group × Trial interaction (F(1,1536) = 4.70, p = 0.030), indicating that RT-scale dynamics evolved differently over trials in SI+ and SI− participants. No main effect of Group was observed, and planned contrasts at trials 1–6 did not survive FDR correction (all q-values > 0.33). In contrast, no Group × Trial interaction was observed in “Life + Me” blocks (all p > 0.55), indicating that group differences in trial-level RT dynamics were specific to the “Death + Me” condition.

Because SI-related dynamics may be expressed as latent temporal structure rather than isolated trial-wise effects, we next examined RT trajectories jointly as multivariate time series. Specifically, we applied a monotonic nonlinear transformation followed by uncentered principal component analysis (PCA) to RT-scale trajectories separately for “Death + Me” and “Life + Me” blocks. The transformation down-weighted large single-trial RT deviations from baseline while preserving the temporal ordering and overall shape of each trajectory, allowing PCA to emphasize reproducible temporal structure rather than isolated outlier trials. Details of the nonlinear transformation and uncentered PCA are provided in the Supplementary Information, **Section 13**. In “Death + Me” blocks, where group-dependent temporal separation was strongest, the first principal component (PC1; **Fig. 3C**) captured an early-trial pattern resembling the divergence observed in the full RT-scale trajectories (**Fig. 3A**). Group differences in PC1 loadings were most pronounced around trial 4 (q ≤ 0.05), and this effect was stable across transformations near the selected parameter range. In contrast, the corresponding PC1 trajectory for “Life + Me” blocks showed weaker and less temporally localized group separation (**Fig. 3D**), consistent with the mixed-effects results. Analyses of higher-order components are provided in Supplementary Information, **Section 14** and **Fig. S4**. To rule out stimulus-specific artifacts, we examined stimulus-word distributions across trial indices and found no evidence of preferential stimulus presentation at early trials (or across SI+/− groups) (Supplementary Information, **Section 15** and **Fig. S5**).

Together, these findings indicate that active SI is reflected in the temporal organization of trial-by-trial RT adjustments, with effects most evident in “Death + Me” blocks. Although both groups showed an initial RT cost followed by rapid recovery, SI+ participants showed a distinct and more variable recovery profile. These results suggest that SI is associated not only with block-level differences or isolated trial effects, but also with latent trial-level structure in task-evoked behavior during decision-making under cognitive control.

### SI State Can Be Robustly Predicted from RT Time Series

Motivated by the group-level temporal effects described above, we next tested whether RT trajectories could distinguish SI+ and SI− participants. We employed a bilinear logistic regression model that incorporates supervised learning of low-dimensional projections of RT dynamics. The model combines both block-level and trial-level mappings and learns multiple latent projections that extract informative features from RTs for classification. We restricted model features to those derived from “Death + Me” blocks, where RT temporal structure was strongest (**Fig. 3C**) and group differences in autocorrelation were most pronounced (**Fig. 2D**). This analytic choice helped maximize the ratio of available data to model degrees of freedom – increasing the tractability of the learning problem and reducing the chances of overfitting and non-convergence. In this framework, the model projects each participant’s “Death + Me” RT matrix onto learned block- and trial-level components, yielding low-dimensional participant-level features for classification (**Fig. 4A**). Details of the bilinear logistic regression classifier are provided in the Supplementary Information, **Section 16**.

**Figure 4.**
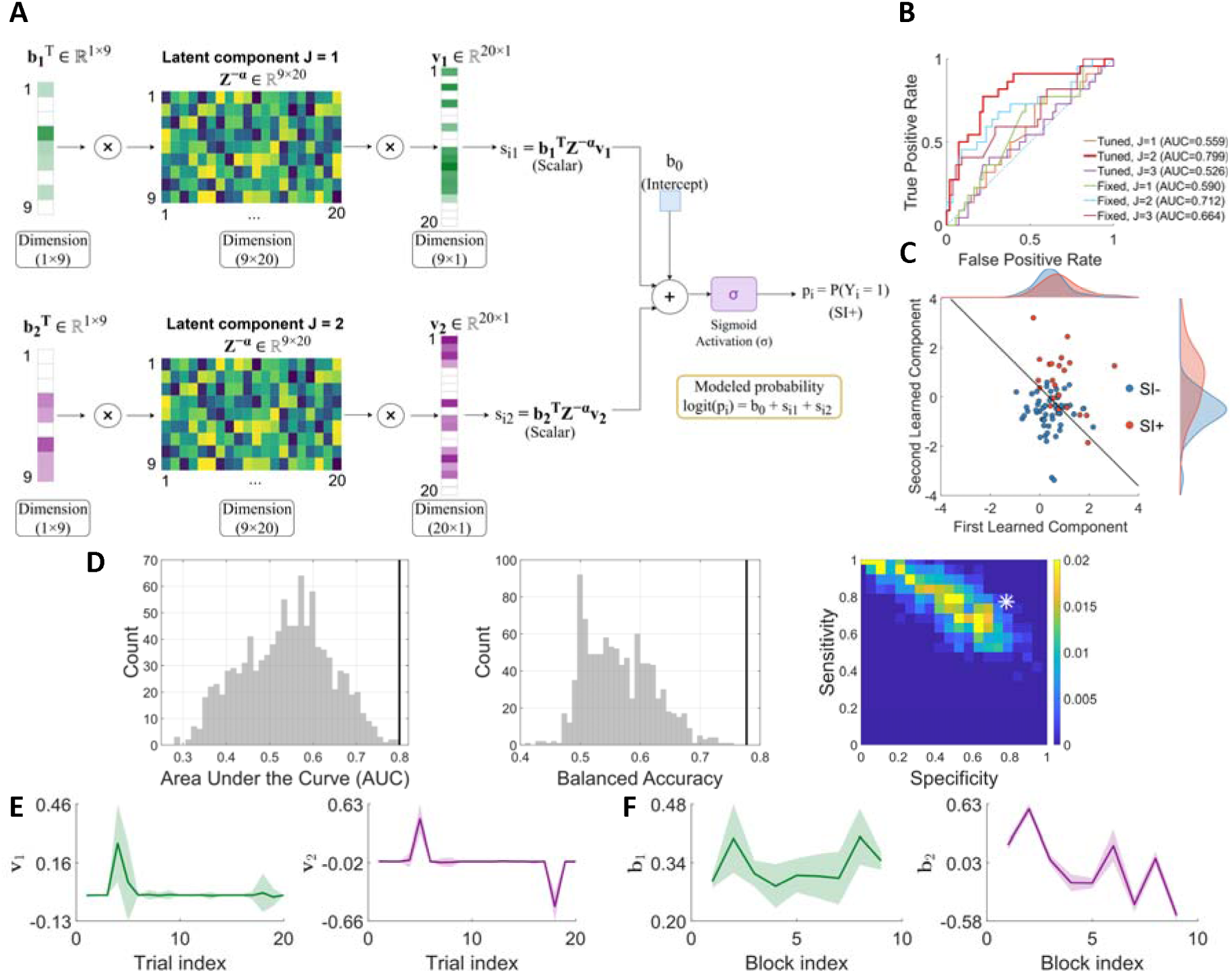
Classification of Suicidal Ideation (SI) State from Spatiotemporal Patterns in Reaction Time (RT) Series. **(A)** Schematic of the bilinear logistic regression classifier used to decode SI state from RT series. For each participant, the trial-by-block RT matrix is projected onto learned temporal weight vectors (***ν_j_***) and block-loading vectors (***b_j_***), yielding low-dimensional features that are passed to a logistic function for classification. In the model shown here, two paired temporal and block components are used to distinguish participants with active suicidal ideation (SI+) from those without active suicidal ideation (SI−). **(B)** Receiver operating characteristic (ROC) curves for classifiers with *J* = 1, 2, and 3 learned spatiotemporal components, evaluated using leave-one-out cross-validation (LOOCV). The model with *J* = 2 achieved the best performance (AUC = 0.799). Baseline models using fixed principal component (PC) directions are shown for comparison and perform less well overall, indicating that SI-relevant structure is better captured by task-optimized bilinear components than by variance-maximizing directions alone. **(C)** Participant-level projections onto the two learned components, shown as 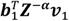 and 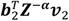, aggregated across LOOCV folds. Points are colored by SI group, and the decision boundary is shown in black. The two-dimensional projection provides partial separation between SI+ and SI− participants, although overlap remains. **(D)** Empirical null distributions obtained from 1,000 random label permutations. **Left**: area under the curve (AUC). **Center**: balanced accuracy. Black vertical lines indicate the observed performance using the true labels. In both cases, the observed values exceed the 95th percentile of the null distribution (*p* < 0.01), indicating that classification performance is unlikely to have arisen by chance. **Right**: Joint null distribution of sensitivity and specificity, with the observed model performance marked by the asterisk (sensitivity = 0.77, specificity = 0.78). **(E)** Learned temporal weight vectors (***ν_j_***), showing the relative contribution of trials within a block. The first component (***ν*_1_**) places strong weight on early trials, particularly around trial 4, whereas the second component (***ν*_2_**) captures a distinct pattern involving both early and later trial positions. **(F)** Learned block-loading vectors (***b_j_***), showing the relative contribution of the nine “Death + Me” blocks. The first component (***b*_1_**) varies modestly across blocks, whereas the second (***b*_2_**) shows stronger block-to-block variation, indicating that SI-relevant temporal structure is not uniform across the task. For **(E)** and **(F)**, lines and shaded bands denote means and 95% bootstrap CIs.

**Fig. 4B** shows ROC curves for classifiers with different numbers of learned components, evaluated using leave-one-out cross-validation (LOOCV). The two-component model achieved the best performance, yielding a balanced accuracy of 77% (specificity = 0.78, sensitivity = 0.77) and an AUC of 0.799. In contrast, one- and three-component models performed substantially worse, with AUCs of 0.559 and 0.526, respectively. **Fig. 4C** shows the joint distribution of the learned projections from the two-component model, indicating that multiple distinct temporal encoding patterns jointly contribute to SI-state prediction. Increasing model complexity to three components appeared to reduce generalization, consistent with overfitting in this modest sample. We also evaluated fixed-PC baseline models in which block-level projections were defined by the leading one or two PCs rather than learned from the classification objective. As shown in **Fig. 4B**, these fixed-PC models achieved AUCs of approximately 0.59 and 0.66, respectively. These results indicate that directions that capture maximal variance in block-level RTs do not reliably correspond to latent projections that optimally encode SI state. Label-shuffling permutation tests confirmed that the observed balanced accuracy and AUC exceeded chance expectations (p < 0.001; **Fig. 4D**).

To rigorously evaluate the proposed model, we benchmarked it against a representative suite of standard machine learning architectures across diverse paradigms, including linear, shallow neural network, recurrent, attention-based, and embedding-based classifiers. These baseline models consistently performed below the proposed bilinear model, underscoring the value of explicitly incorporating task-informed temporal structure into the feature set. Details of model configurations and performance comparisons are provided in Supplementary Information, **Section 17**.

**Figs. 4E-F** show the learned temporal weight vectors and block-loading vectors for the best-performing two-component model. The first temporal component places its strongest weights on early trials within each block, whereas the corresponding block-loading vector exhibits modest elevations during the initial and final blocks but otherwise maintains a consistent baseline. This pattern is consistent with a dominant, block-invariant temporal encoding component and aligns with the principal temporal structure identified in the autocorrelation and PCA analyses (**Fig. 3C**). By contrast, the second component exhibits a more heterogeneous pattern: trial weights included both positive and negative values, and block loadings varied more strongly across the task, suggesting that the second component captures a higher-order temporal pattern whose expression depends more strongly on task stage and may reflect time-dependent processes, such as adaptation or fatigue. Projections onto the second component (**Fig. 4C**) revealed a more restricted distribution in SI− participants, whereas SI+ participants showed broader dispersion in this latent space. This pattern suggests that the block-dependent temporal dynamics captured by the second component are more constrained in the SI− group and more heterogeneous in the SI+ group.

Together, these analyses show that temporal RT dynamics identified at the group level also support participant-level prediction of active SI through directly inspectable block- and trial-level features. Given the nature of our primary sample, one might wonder whether our model simply learned to classify people by SI-associated features – such as MDD diagnostic status – rather than by SI itself. To assess this, we reapplied the same bilinear classification pipeline using major depressive disorder (MDD) versus control labels instead of EMA-derived SI labels. Classification performance was reduced, with AUC decreasing from 0.799 for SI classification to 0.601 for MDD-control classification (Supplementary Information, **Section 18**). These results suggest that the RT dynamics captured by the model are more informative about near-term SI state than about MDD diagnostic status in this cohort (i.e., that our model did not simply learn to classify people on diagnostic status).

## Discussion

Our results demonstrate that near-term active suicidal ideation is reflected in the temporal dynamics of RTs during a modified BDIAT. Information on active SI could be gleaned from analyses of these dynamics at several temporal scales, including block-to-block transitions and fine-grained trial-level dynamics within blocks. Critically, these dynamics were more informative than conventional D-scores, which were unable to distinguish SI+ from SI− participants.

The limited informativeness of the D-score in this study contrasts with prior work showing modest predictive value of death/suicide IAT scores for suicide-related histories and risk groups(10,11). This difference may result from our use of EMA to ascertain SI status. EMA captures short-term experiences in daily life, which is a departure from previous studies’ use of interview-based measures and their focus on long-standing histories of suicidality. The (modest) informativeness of the D-score in those studies, together with their lack of informativeness here, might lead one to conclude that the D-score reflects relatively enduring or structurally embedded circuit abnormalities associated with suicide attempts or chronic high-risk states. This conclusion accords with the fact that changes in amygdala–prefrontal connectivity and salience-network organization have been linked to stronger suicidal self-associations on implicit tasks and to elevated risk for attempts(31). It also aligns with the fact that D-scores do not change alongside ketamine-induced reductions in suicidality(32). In contrast, trial-by-trial RT dynamics may capture more transient alterations in cognitive control, conflict adaptation, and contextual sensitivity that contribute to the near-term SI measured here by EMA. Because these processes unfold during task performance, they may be more sensitive to current clinical state and therefore better positioned to track change over short timescales(33,34). Thus, temporal BDIAT features may complement, rather than replace, conventional D-scores by targeting a more state-like dimension of suicide risk.

Our study identified these SI-associated temporal BDIAT features across two distinct analysis windows. At the block level, autocorrelation analyses revealed a rhythmic RT pattern locked to the alternating BDIAT block structure. SI− participants showed stronger and more persistent alignment to block alternation, whereas SI+ participants showed attenuated entrainment. This rhythmic structure generalized to an independent online task variant with shorter blocks, even when group-level discrimination was reduced. This loss of group discrimination may reflect, in part, differences in SI labeling, with the primary cohort using repeated EMA-derived labels designed to capture near-term fluctuations in SI and the online cohort using single-time-point self-report C-SSRS grouping. This interpretation supports our use of EMA-derived labels for our main cohort. However, differences in task setting, administration, and block length may also have contributed. The online setting in particular may have influenced RTs (e.g., because we did not control distractions in the environment as we did in the lab) in ways that add noise and obscure systematic SI-associated differences. Regardless of the exact driver of reduced group-level discrimination, these findings indicate that sensitivity to block-level context is a robust feature of BDIAT performance and that SI may be associated with weaker or less stable encoding of task context across block transitions.

At the trial level, the observed RT dynamics were consistent with altered conflict adaptation in people with SI(16,35). Following block switches, both groups exhibited an initial RT cost, but SI− participants showed more regular recovery toward baseline, whereas SI+ participants showed faster habituation and greater variability. These differences were modest on individual trials but became clearer when RT trajectories were analyzed as multivariate time series. Low-dimensional temporal representations captured early post-switch dynamics, suggesting that near-term SI is associated with altered context updating rather than uniform slowing or generalized performance deficits. The broader dispersion of SI+ participants in classifier space (**Fig. 4C**) is consistent with the block-level autocorrelation findings (**Figs. 2E-F**) and suggests that SI is associated less with a uniform directional shift in behavior than with greater heterogeneity in task-evoked temporal dynamics. This heterogeneity may help explain why aggregate measures such as D-scores, which reduce performance to a single summary value, fail to reliably distinguish SI groups. Future studies might examine whether this heterogeneity in BDIAT RT dynamics is meaningfully linked to heterogeneity in the underlying causes of an individual’s suicidality. If so, our approach may have applications in personalized medicine.

Importantly, our models avoid a common pitfall of machine learning studies of suicidality. It is easy to accidently build models that distinguish people according to group features, such as mental health diagnosis, that correlate with SI, instead of models that identify SI per se. Our models are not likely to have done this. Diagnostic-label analyses suggested that the temporal features used in these models were more closely related to near-term SI than to broad MDD versus control status (Supplementary Information, **Section 18**). Our study also explicitly demonstrates that taking into account temporal BDIAT structures carries information about suicidality above and beyond that contained in other features of the task. This is implied by the non-informativeness of the D-score. It is more rigorously shown by the fact that alternative classifiers that did not explicitly incorporate task-informed temporal structure performed worse than the proposed bilinear model (Supplementary Information, **Section 17**). Together, these findings support the value of compact, interpretable models that preserve the temporal organization of BDIAT behavior while limiting overfitting in modest clinical samples.

Several limitations should be acknowledged. First, SI state was binarized across the EMA period, which made classification tractable but likely obscured heterogeneity and fine-grained differences in SI severity, frequency, and phenomenology within the SI+ group. Second, the modest sample size required leave-one-out cross-validation and precluded a separate independent test set; larger cohorts will be needed to establish generalizability. Because suicidality is a transdiagnostic phenomenon, these cohorts should ideally include individuals from multiple groups at high-risk for self-harm, such as people with psychosis or with anorexia. Third, because the current analyses paired one behavioral session with one set of EMA-derived labels, without repetition of this procedure, we could not determine whether these temporal features track within-person changes in suicidality or treatment response. Future longitudinal work should test whether these signatures change with or auger changes in suicidality and whether they are strengthened by integration with additional modalities, ideally without loss of scalability or interpretability.

Indeed, a key advantage of the current behavior-based approach is its simplicity and scalability. Because the task requires only a standard computer or smartphone, it could support repeated, low-burden assessment in settings with limited access to specialized clinical resources (e.g., neuroimaging technologies). Accordingly, this framework may offer a practical route toward more accessible and timely evaluation of near-term suicide risk that is more compatible with known rates of fluctuation in SI. Because temporal RT dynamics are not a widely known marker of suicidality, and because these dynamics are complex and thus difficult to spoof, our approach may also be a more robust indicator of SI than self-report, which can be easily altered if one desires to hide suicidality. With further validation in larger and longitudinal cohorts, our approach could eventually support targeted intervention and treatment monitoring in real-world settings.

## Data availability

The data supporting the findings of this study contain sensitive clinical and behavioral information from human participants and are therefore not publicly available. De-identified data may be made available from the corresponding author upon reasonable request, subject to institutional review board (IRB) approval and applicable data-use agreements.

## Code availability

All analyses were conducted in MATLAB and Python using the Statistics and Machine Learning Toolbox and the lme4/mixedlm implementations. Code for all analyses and figure generation is publicly available at https://github.com/YousefiNeuroLab/Near-Term-Suicidality-Risk-Encoded-in-the-Temporal-Dynamics-of-the-Death-IAT.

## Acknowledgments

This project is sponsored by the Defense Advanced Research Projects Agency (DARPA) under cooperative agreement No. N660012324016. The content of the information does not necessarily reflect the position or the policy of the Government, and no official endorsement should be inferred. This work was supported by the Defense Advanced Research Projects Agency (grant number N660012324016; A.Y., M.V.B., A.S.W.).

## AI Tool Use Statement

During manuscript preparation, the authors used AI-based language models and grammar-checking tools to improve readability, language clarity, and basic formatting. These tools were not used to generate, analyze, or interpret scientific data, nor to draw scientific conclusions; all analyses, results, and interpretations are the sole work and responsibility of the authors.

## Ethical Considerations

The authors assert that all procedures contributing to this work comply with the ethical standards of the relevant national and institutional committees on human experimentation and with the Helsinki Declaration of 1975, as revised in 2008. The study was approved by the Advarra Institutional Review Board (IRB), with oversight from the University of Minnesota and collaborating institutions. All participants provided written informed consent and HIPAA authorization under a protocol approved by the Advarra IRB and the U.S. Navy Human Research Protection Office (HRPO). Additional safeguards were implemented for participants expressing acute suicidal intent, consistent with predefined clinical response procedures.

## Author Contributions

P.R., A.S.W., and A.Y. contributed to manuscript preparation, data analysis, and algorithm development. M.V.B. designed major components of the task, oversaw experimental protocols, and assisted with manuscript preparation. B.M.G., S.Y., A.N.M., B.B., M.F., and S.M.B. contributed to patient recruitment, data recording, and data cleaning. S.T. contributed to data analysis. M.D.G.K. contributed to IRB protocol development and project management.

