## Supplementary Information for "Predicting Active Suicidality from Temporal Dynamics of Implicit Life-Death Associations"

**Supporting Information Text**

**Section 1. Participants.** Participants were recruited as part of a larger research effort focused on identifying neurobehavioral markers of suicide risk. To be generally eligible, participants needed to be age 18-65, be fluent in English, and be able to read computer-presented text (with corrective lenses if necessary). Exclusion criteria included neurological disorders, contraindications to electroencephalography (EEG) (e.g., scalp sensitivity or gel allergy), pregnancy, recent psychiatric hospitalization, or active military or Department of Defense employment (due to funders’ requirements). Participants with treatment-resistant depression and histories of suicidality who met these criteria were recruited from interventional psychiatry clinics. These participants were required to have planned treatment with transcranial magnetic stimulation (TMS) or ketamine, to support potential analyses focused on clinical change. A comparison group consisting of community members, who did not have any psychiatric condition and were therefore less likely to report suicidality, were recruited via StudyFinder. All participants provided written informed consent under a protocol approved by the Advarra Institutional Review Board (IRB) and the U.S. Navy Human Research Protection Office (HRPO) under Defense Advanced Research Projects Agency (DARPA) cooperative agreement No. N660012324016.

After screening for data quality (e.g., participants were excluded for BDIAT accuracies <10%, indicating instructions were not understood and followed) and protocol adherence (e.g., completion of the BDIAT and EMAs), the eligible sample included n=55 people with depression and n=25 comparison group members.

**Section 2. Participant Enrollment and Data Quality Exclusions**. Participant enrollment and data quality exclusions. Data quality thresholds were applied before preprocessing in both cohorts. Trials with RT < 350 ms or RT > 2,000 ms were treated as invalid, and participants with more than 10% missing or invalid trials were excluded from downstream analyses.

In the primary in-person cohort, 80 participants completed EMA and the BDIAT. Based on EMA responses, 57 were classified as SI− and 23 as SI+. Application of the data quality threshold excluded 3 participants: 2 from the SI− group and 1 from the SI+ group. The final primary analytic sample included 77 participants (SI− = 55, SI+ = 22). In the online validation cohort, 100 participants were enrolled through Prolific, 94 completed the task, and 11 were excluded based on the same data quality threshold. The final online validation sample included 83 participants. The enrollment, allocation, and exclusion flow for both cohorts are shown in **Fig. S1.**

**Figure S1. Flowchart of Participant Enrollment and Data Quality Exclusions.** Flowcharts show participant inclusion, group assignment, data quality exclusions, and final analytic samples for the primary and online validation cohorts. **(A)** In the primary cohort, 80 participants completed EMA and the BDIAT and were classified by EMA-derived SI status as SI− (n = 57) or SI+ (n = 23). Applying the data quality threshold (>10% missing or invalid trials; invalid RT defined as RT < 350 ms or RT > 2,000 ms) excluded 2 SI− participants and 1 SI+ participant, yielding a final analytic sample of 77 participants (SI− = 55, SI+ = 22). **(B)** In the online validation cohort, 100 participants were enrolled through Prolific and 94 completed the task. Applying the same data quality threshold excluded 11 participants, yielding a final online validation sample of 83 participants.

.

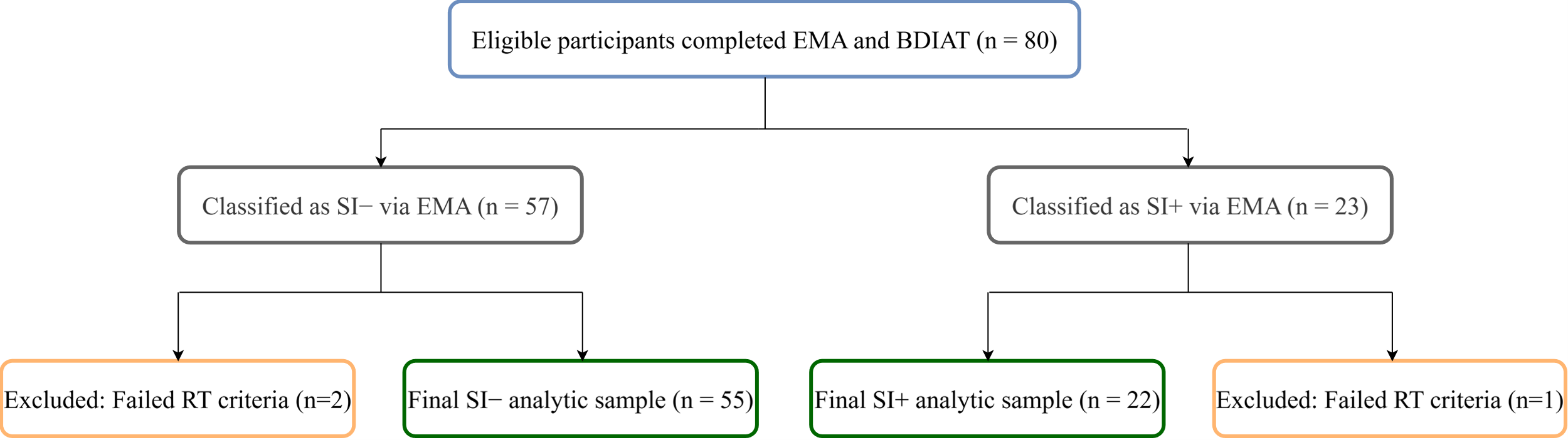

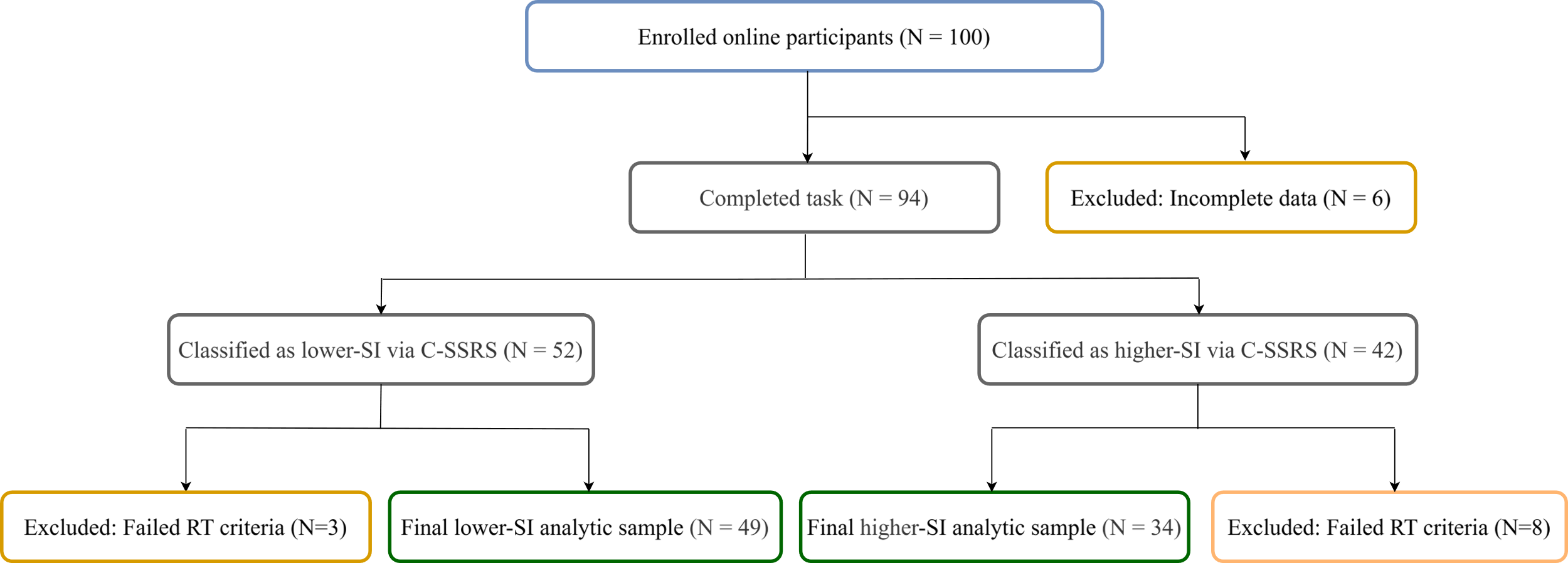

A

B

**Section 3. EMA and Clinical Measures.** To obtain a repeated, near-term measure of SI in daily life, participants completed EMA surveys delivered to their smartphones three times per day for seven consecutive days prior to the BDIAT session. EMA has been shown to capture short-term fluctuations in suicidal thoughts and reduce recall bias compared to retrospective clinical assessment(1–5). Each survey included items assessing passive suicidal ideation (e.g., “I wish I were not alive”), active suicidal ideation (e.g., “I am thinking about killing myself”), and theoretically relevant contributors to such ideation, including psychological pain, all of which were rated on a 0–4 scale (0 = not at all, 4 = extremely). Participants were classified as SI+ if their mean EMA active SI score across completed prompts was greater than zero, indicating at least one endorsement of active suicidal thoughts during the monitoring period(4,6). Participants who scored zero on active SI items across all completed EMA prompts were classified as SI-. This binary EMA-derived label was used as the primary outcome variable for all classification models, corresponding to the SI+ and SI− groups.

Among the 80 eligible participants, 57 were assigned to the SI− group and 23 to the SI+ group based on EMA responses. Three participants were subsequently excluded because their BDIAT sessions exceeded the prespecified threshold for missing data (>10% missing trials): two from the SI− group and one from the SI+ group. This yielded a final analytic sample of 77 participants (55 SI− and 22 SI+), for whom the EMA-derived SI label was used as the primary outcome variable in all behavioral and machine learning analyses.

In addition to EMA, participants completed the C-SSRS(7) during the laboratory visit. The C-SSRS was administered by trained research assistants. The C-SSRS assesses recent and lifetime suicidal ideation, intent, and behavior. However, consistent with prior work, several participants ($n=10$) who did not endorse active SI on the C-SSRS reported active suicidal thoughts on EMAs. In contrast, only two people were identified as having suicidal thoughts on the C-SSRS who were not detected by EMAs. Taken together, these results suggest that single timepoint assessments, like the C-SSRS, may miss transient ideation captured by repeated real-time self-report(2,8), whereas dense measurement strategies are comparatively unlikely to miss ideators. With this in mind, C-SSRS scores were used only as a secondary clinical benchmark, whereas the EMA-derived active SI label served as the primary indicator of near-term active ideation.

**Section 4 Behavioral Task: Brief Death Implicit Association Test.** The BDIAT, which measures implicit associations between the self and life or death, was administered approximately one week after the EMA period. The task consisted of 360 trials, organized into 18 alternating blocks: 9 “Life + Me” and 9 “Death + Me” blocks, with 20 trials per block (**Fig. 1B**). The category pair for each block remained on-screen throughout that block. In each trial, participants sorted stimuli as quickly and accurately as possible according to whether the presented word matched either category label. Stimulus response mappings alternated between blocks accordingly with the alternating category labels, and the assignment of response keys (left/right) was counterbalanced across participants to minimize motor biases. By alternating between “Life + Me” and “Death + Me” blocks, the task was designed to capture variation in RT during self-referential life/death categorization.

On each trial, a fixation cross appeared for $500 \pm50 ms$, followed by presentation of a single word stimulus at the center of the screen. Stimuli included self-related words (e.g., me, myself, I), other-related words (e.g., they, them, other), death-related words (e.g., dead, suicide, lifeless, buried), and life-related words (e.g., alive, living, breathe, survive). The word remained on screen for up to 2 seconds or until a response was made. To ensure participants understood the lexical meaning of each stimulus word, prior to beginning experimental trials, participants completed a word sorting task in which they sorted the task stimuli into Death/Life/Me/Other categories. Following the word sorting task, participants completed 10 practice trials of the BDIAT task, in which feedback regarding correct/incorrect responses was displayed at the end of each trial. No feedback was displayed in experimental trials.

Reaction times and accuracy were recorded for every trial, which translates to 360 trials per participant. The full task lasted approximately 12–14 minutes. In this study, BDIAT performance served as the primary behavioral measure to investigate whether temporal dynamics of RTs could reveal markers of acute suicidal ideation.

**Section 5. Online BDIAT validation cohort.** To assess whether block-level rhythmicity generalized across task parameters and data-collection settings, we analyzed an independent online BDIAT cohort. Participants were recruited through Prolific and completed an online BDIAT variant consisting of 36 alternating blocks with 10 trials per block. This is half as many trials per block as the in-person sample, which we chose to verify that our observed temporal patterning was robust to small changes in task design. Of 100 enrolled participants, 94 completed the task; 83 were retained after excluding 11 participants with an excessive proportion (>10%) of missing or invalid trials (defined as RT < 350 ms or > 2,000 ms). Participants were grouped into lower- and higher-SI categories using self-reported C-SSRS responses, with any nonzero endorsement of suicidal ideation (C-SSRS > 0) classified as higher-SI (n=34) and no endorsement (C-SSRS = 0) classified as lower-SI (n=49); this any-versus-none cutoff was chosen to mirror the binarization rule applied to the EMA-derived SI label in the primary cohort, preserving a comparable SI definition across cohorts. This grouping approximated the SI− versus SI+ contrast used in the EMA-characterized cohort, but differed in assessment instrument, temporal resolution, and testing environment.

**Section 6. D-Score Definition and Computation.** The D-score, a commonly used behavioral index of implicit association strength, was defined here as the standardized difference in mean RT between the “Death + Me” and “Life + Me” blocks. Larger D-scores indicated relatively slower responses in “Death + Me” blocks than in “Life + Me” blocks. The D-score was calculated as follows:

$D=\frac{\mu_{d}-\mu_{l}}{\sigma_{\text{all}\text{ }}},$	(1)

where $\mu_{d}$ is the mean RT for trials in “Death + Me” blocks, $\mu_{l}$ is the mean RT for trials in “Life + Me” blocks, and $\sigma_{all}$ is the pooled standard deviation of RTs across both block types(9,10). Before computing D-scores, RT data were cleaned to reduce the influence of invalid trials. Trials with excessively fast responses (RT < 350 ms) and trials with no response within the response window (RT > 2000 ms) were excluded from the mean RT calculations. D-scores were then computed using the improved IAT scoring algorithm(9). This approach uses the pooled standard deviation to standardize block-wise RT differences and is commonly used in the IAT literature because it reduces the influence of individual differences in overall response speed(9,11).

**Section 7. Preprocessing and Normalization.** RT sequences were preprocessed to address skewness, censoring, and missing or physiologically implausible responses prior to temporal analysis. For each participant $i$, we denote the raw RT sequence as

$\mathbf{r}_{i}=\{r_{it}{\}}_{t=1}^{T},$	(2)

where $T$ is the total number of trials. Trials with no recorded response were treated as right-censored observations (RT exceeding a fixed upper bound of 2,000 ms), while extremely fast responses below a conservative threshold of 350 ms were considered physiologically implausible. These trials were excluded from direct analysis.

Following standard practice for reaction time data, RTs were log-transformed to stabilize variance and reduce skewness. Let $y_{it}$ denote the transformed observation, defined as

$y_{it}=\left\{ \begin{matrix} \text{censored}, & r_{it}>\tau_{\text{max}}, \\ \text{missing}, & r_{it}<\tau_{\text{min}}, \\ \log(r_{it}), & \text{otherwise}, \end{matrix} \right.$	(3)

where $\tau_{\text{max}}$ and $\tau_{\text{min}}$ denote upper (2,000 ms) and lower (350 ms) RT thresholds, respectively. RT data can be preprocessed in multiple ways, each with their own strengths and limitations. Because RT sequences in this task include censoring, physiologically implausible responses, and temporal non-stationarity, we evaluated two complementary preprocessing strategies to ensure that our findings were not dependent on a particular modeling assumption. The first approach uses a state-space formulation that explicitly accounts for censoring and missingness within a probabilistic framework and assumption of a state-like RT(12). The second approach applies a threshold-aware local imputation procedure designed to preserve short-range temporal structure with minimal model assumptions. The state-space method served as our primary preprocessing pipeline, while the local imputation approach was used as a robustness check. Across analyses, results were qualitatively consistent across preprocessing strategies (Supplementary Information, **Section 8**).

Our primary analyses were conducted using the state-space formulation, where we modeled the log-transformed RT sequence using a state-space formulation that explicitly accounts for censoring and missing data. Specifically, the observed signal was assumed to arise from a latent dynamic process,

$$\begin{aligned} y_{it}=c_{i}+x_{it}+\epsilon_{it}, \epsilon_{it}\mathcal{\sim N}\left( 0,\sigma_{y,i}^{2} \right),\#\left( 4 \right) \end{aligned}$$

with latent state evolution

$$\begin{aligned} x_{it}=x_{i,t-1}+v_{it},v_{it}\mathcal{\sim N}\left( 0,\sigma_{x}^{2} \right),\#\left( 5 \right) \end{aligned}$$

and prior $x_{i0}\mathcal{\sim N}(0,\sigma_{0}^{2})$. Here, $c_{i}$ represents a participant-specific baseline, $\sigma_{x}^{2}$ controls the smoothness of the latent trajectory, and $\sigma_{0}^{2}$ is set to a large, fixed value to reflect weak prior information. Censored observations contributed to the likelihood through their censoring bounds, while missing observations were handled using a flat likelihood. Model fitting was performed independently for each participant using a Bayesian filtering approach implemented in the COMPASS toolbox, as described previously(13). The posterior mean of the latent process, $x_{k}$, represents the smoothed log-reaction time trajectory. This latent state was subsequently transformed into RT-scale, which represents reaction time as a multiplicative deviation from the participant-specific baseline, as detailed below.

As an alternative preprocessing strategy, missing values in the log-transformed RT sequences were imputed using a threshold-aware local estimation procedure. In this approach, each missing value was replaced using statistics computed from a local temporal window centered on the missing trial (±15 trials, truncated at the series boundaries), with censoring information used to modulate the imputation rule. An upper bound corresponding to the log-transformed task response limit was applied during imputation to prevent unrealistically large values for censored trials. This procedure preserves local temporal structure while limiting the influence of extreme or censored observations. The resulting imputed log-transformed RT sequences were used for all downstream temporal analyses and modeling.

For preprocessing and filtering, the initial state variance was set to $\sigma_{0}^{2}=1$, and the state noise variance was set to $\sigma_{x}^{2}=0.12$. RTs below $t_{\min}=350$ ms were treated as physiologically implausible given the speed with which visual information enters conscious awareness, and trials exceeding the response window ($t_{\max}=2000$ ms) were treated as right-censored. Within the state-space formulation, participant-specific baseline offsets $c_{i}$ and observation noise variances $\sigma_{y,i}^{2}$ were estimated from the data to achieve an optimal fit for each participant.

We exponentiated the posterior mean of the latent state $\boldsymbol{x}_{i}\boldsymbol{=}\{x_{it}{\}}_{t=1}^{T}$ **to obtain the RT-scale signal** $\boldsymbol{z}_{i}$**, where** $z_{it}\boldsymbol{=}\exp\left( x_{it} \right)$**.**

$$\begin{aligned} RTscale_{k}=\exp\left( x_{k} \right)\#\left( 6 \right) \end{aligned}$$

**This quantity specifies how each trial is scaled multiplicatively relative to the participant-specific baseline RT. Alternative parameter settings and preprocessing variants were examined to assess robustness, and results were consistent across choices.**

Participants with an excessive proportion (>10%) of missing or invalid trials were excluded prior to preprocessing to ensure reliable estimation of temporal dynamics. Invalid trials were defined as responses with RT < 350 ms or RT > 2,000 ms. Specifically, three participants were excluded from the in-person cohort, and 11 participants were excluded from the online validation cohort (Supplementary Information, **Section 2**).

**Section 8. Robustness of Temporal Features to Preprocessing and Imputation Strategies.** The primary analyses reported in the main text used a state-space formulation to infer continuous latent RT trajectories while accounting for missing and censored observations. To verify that the temporal features and classification results were not dependent on this preprocessing choice, we repeated the analysis using an alternative threshold-aware local imputation procedure.

In the alternative approach, missing values in the log-transformed RT sequence were imputed using statistics computed from a local temporal window centered on the missing trial (±15 trials, truncated at the sequence boundaries). Censored trials were constrained using an upper bound corresponding to the log-transformed task response limit. Representative participants with missing and censored trials showed highly similar temporal structure under the primary state-space filter and the local imputation procedure (**Fig. S2A–B**).

Across all participants and trials, the two preprocessing pipelines produced strongly concordant trajectory estimates (**Fig. S2C**; R² = 0.9878). Small deviations from the identity line were expected because the two approaches make different assumptions about missing and censored observations: the state-space model estimates a smoothed posterior trajectory under a probabilistic censoring model, whereas the local imputation approach estimates missing values from neighboring observations within a fixed temporal window. Thus, discrepancies were most apparent near censored or dynamically variable trials but were limited relative to the overall agreement between pipelines.

To quantify the impact of preprocessing choice on predictive performance, we repeated the participant-level LOOCV analysis using the best-performing bilinear classifier configuration from the primary analysis (J = 2). Classification performance remained comparable under the alternative preprocessing pipeline, with AUC = 0.791 compared with AUC = 0.799 for the primary state-space approach (**Fig. S2D**). Together, these findings indicate that the temporal dynamics supporting SI classification are robust to the preprocessing strategy and are not artifacts of the missing-data handling procedure.

**Figure S2. Robustness of Reaction Time (RT) Dynamics and Classification Performance Across Preprocessing Strategies.** **(A)** Representative log-transformed RT trajectory for a participant with missing and censored trials. Raw log-transformed RTs are shown in black, and the primary state-space posterior mean is shown in teal. Orange crosses mark missing RT trials, blue circles mark censored RT trials, and dashed horizontal lines indicate the lower and upper RT thresholds. **(B)** Same participant as in **(A)**, showing the alternative threshold-aware local imputation trajectory in pink alongside the raw log-transformed RTs. **(C)** Trial-level correspondence between trajectory estimates from the primary state-space pipeline and the secondary local imputation pipeline across all participants and trials. Points are concentrated around the identity line, indicating strong agreement between preprocessing strategies (R² = 0.9878), with modest deviations for a subset of trials. **(D)** ROC curves for the bilinear logistic regression classifier with two learned components (J = 2), evaluated using participant-level leave-one-out cross-validation (LOOCV). Classification performance was similar across preprocessing strategies, with AUC = 0.799 for the primary state-space approach and AUC = 0.791 for the secondary local imputation approach.

A

B

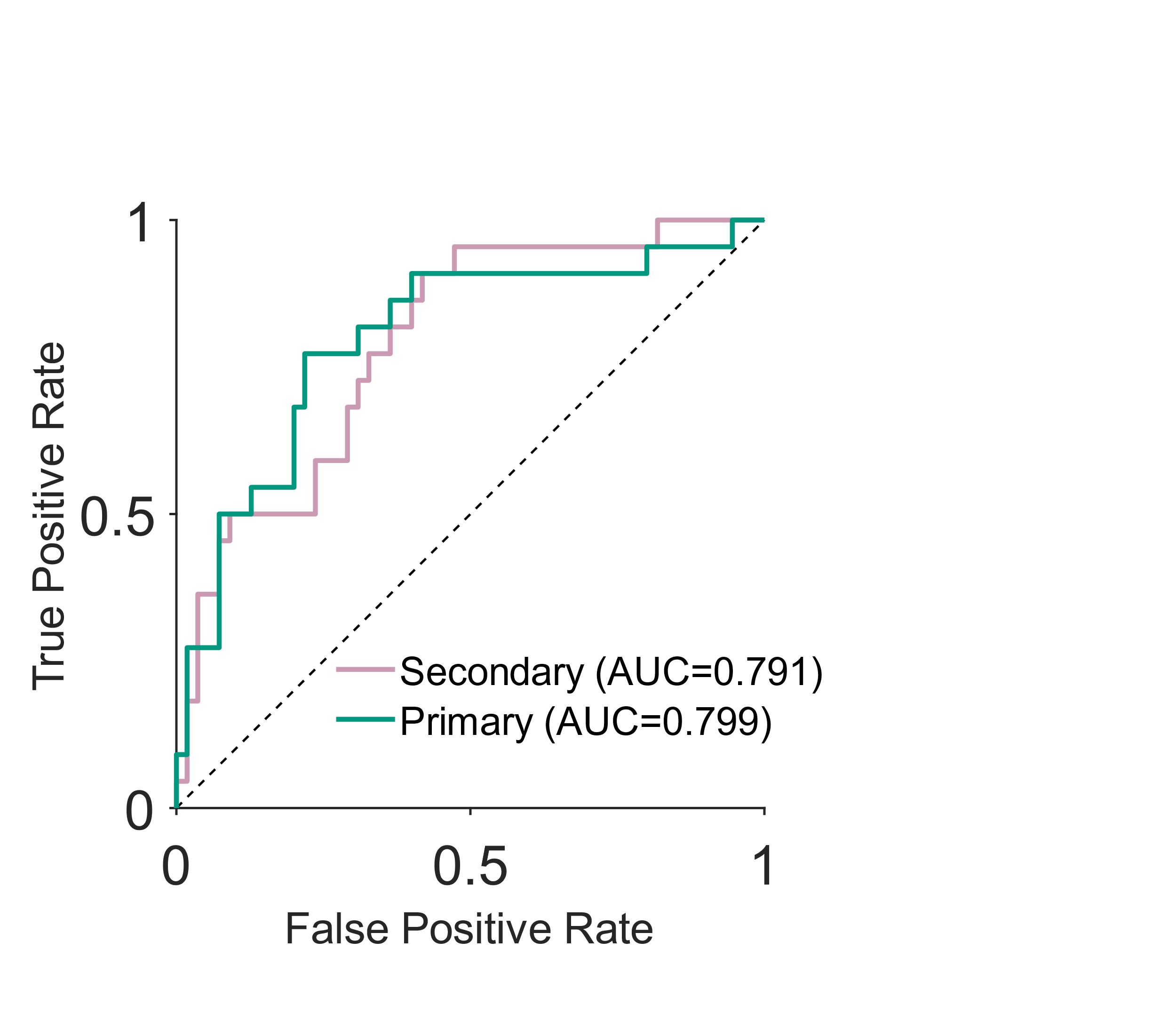

C

D

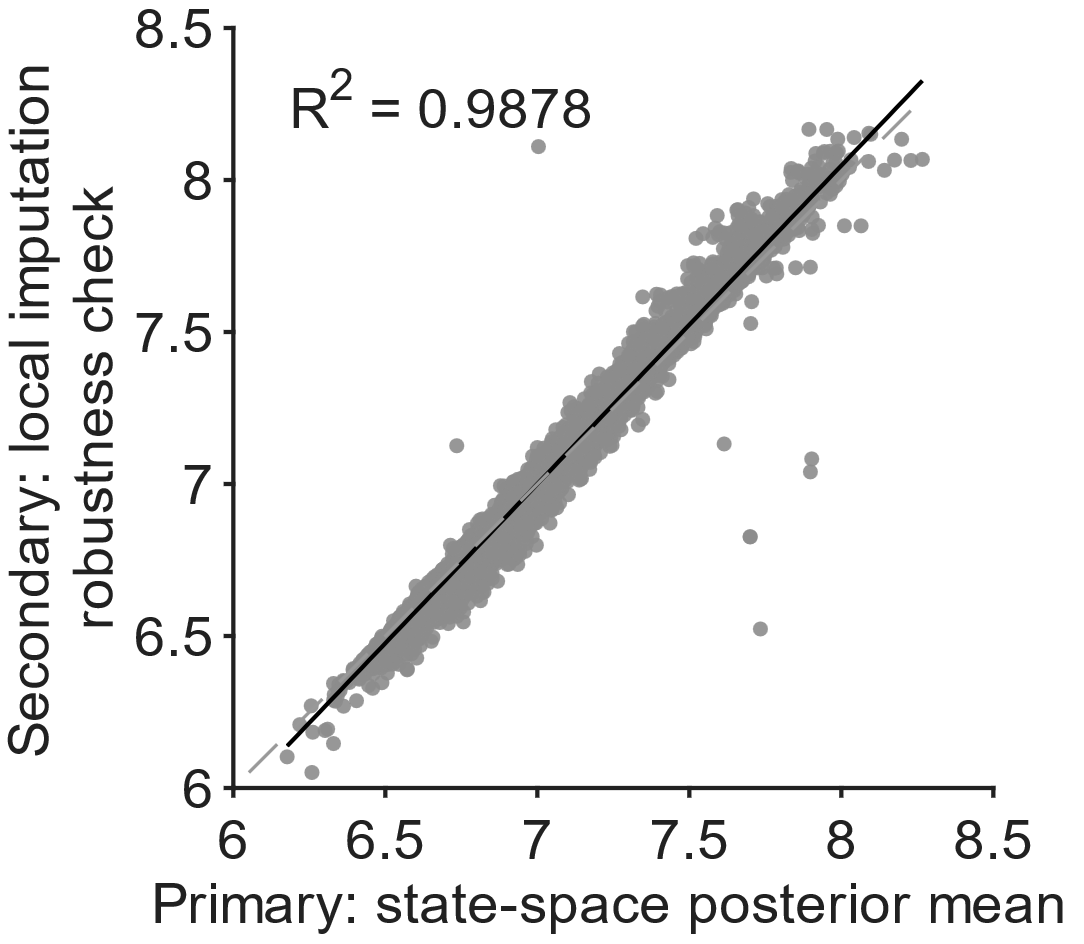

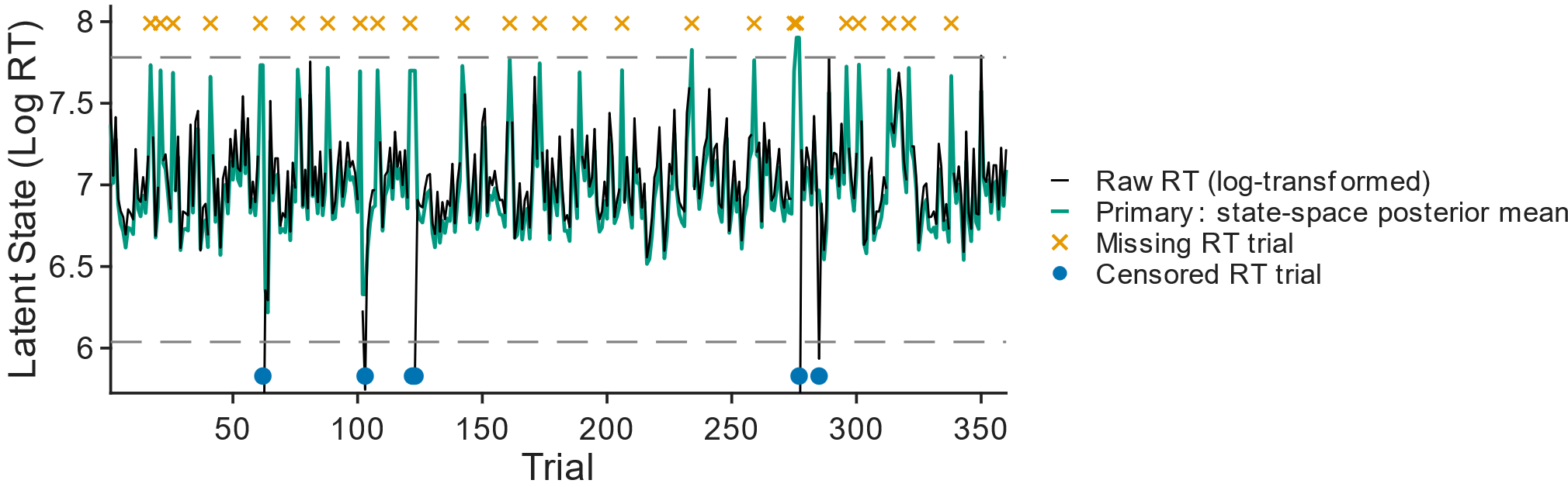

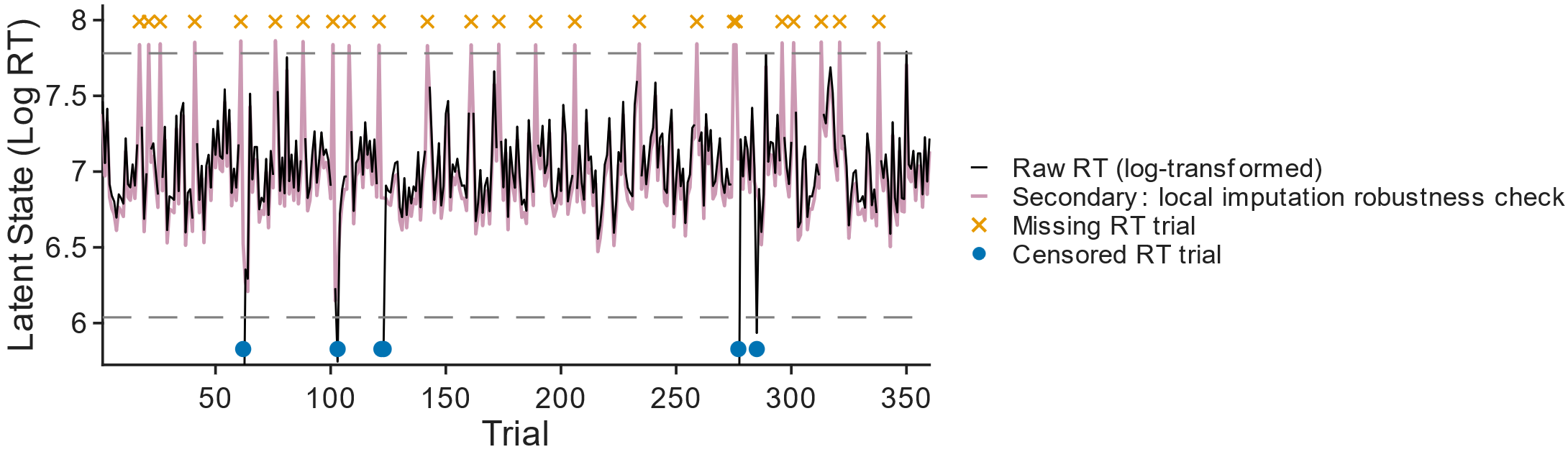

**Section 9. Block-level autocorrelation and Rhythm Index.** Standard scoring frameworks for the BDIAT, such as the D-score, aggregate reaction times across alternating block types (e.g., “Death + Me” vs. “Life + Me”), inherently discarding temporal dynamics and transient conflict processing at block transitions. In preliminary investigations utilizing a smaller sample (27 participants; 18 SI−, 9 SI+)(14), spectral analysis of trial-level RTs revealed enhanced power at the block-switching frequency, a signature that was more pronounced in SI− participants and was associated with more temporally regular patterns of RT fluctuation across blocks. However, frequency-domain approaches are susceptible to spectral leakage when applied to short behavioral time series or when the series does not contain complete cycles of the underlying rhythm. This leakage artifactually distributes power from true oscillatory components into adjacent frequency bins, obscuring precise rhythmic peaks and reducing confidence in distinguishing genuine task-locked structures from sampling artifacts. To robustly isolate temporal dependencies aligned with the task's alternating architecture, we developed an autocorrelation-based pipeline.

For each participant, baseline-scaled RTs were averaged across the 20 trials within each block to form an 18-point block-level time series in the primary cohort, and across the 10 trials within each block to form a 36-point block-level series in the online cohort. Before autocorrelation, block-level series were mean-centered and linearly detrended within participant. We computed autocorrelation functions across lags 1–8. Because the task alternates between “Death + Me” and “Life + Me” blocks, positive autocorrelation at even lags and negative autocorrelation at odd lags indicate alignment with the alternating task structure. We summarized this alignment with a multi-lag Rhythm Index, defined as the mean autocorrelation at even lags minus the mean autocorrelation at odd lags.

To test whether rhythmicity depended on task structure rather than nonspecific temporal dependencies, we compared observed Rhythm Index values with within-subject shuffle nulls. The destroy-ABAB shuffle randomly permuted all blocks, disrupting the alternating order. The preserve-ABAB shuffle permuted blocks within odd and even positions separately, preserving broad alternation while disrupting the original ordering within each task category. Group-level and participant-level shuffle tests are reported in the Results, Rhythmic Entrainment to Block Structure Is Present in BDIAT and Differs Across SI Groups; and Supplementary Information, **Section 10**.

**Section 10. Shuffle-based Evaluation of the Rhythm Index.** To determine whether the block-level rhythmic structure reflected task-locked organization rather than nonspecific temporal dependence in reaction times (RTs), we compared the observed Rhythm Index with within-subject shuffle-derived null distributions. For each participant, block-level RT-scale values were summarized by averaging across trials within each block, then detrended and mean-centered before computing the block-level autocorrelation function. The multi-lag Rhythm Index was defined as the mean autocorrelation at even lags minus the mean autocorrelation at odd lags across lags 1–8.

We generated two within-subject null models. In the destroy-ABAB shuffle, all blocks were randomly permuted, disrupting the alternating task structure while preserving each participant’s set of block-level RT values. In the preserve-ABAB shuffle, blocks from odd positions were permuted only among odd positions and blocks from even positions only among even positions. This preserved the broad ABAB alternation while disrupting the original ordering of blocks within each task category. For each shuffle condition, we generated null distributions of Rhythm Index values across 10,000 shuffles for each participant and computed the difference between the observed Rhythm Index and the median of the corresponding null distribution.

**Fig. S3A–B** shows the observed-minus-null Rhythm Index differences for the primary cohort. In both shuffle conditions, the distributions were significantly shifted above zero. The observed Rhythm Index was significantly greater than the destroy-ABAB null distribution, indicating that the rhythmic structure depended on the alternating organization of task blocks. The observed Rhythm Index also remained significantly elevated relative to the preserve-ABAB null distribution, indicating that the rhythmic structure could not be explained solely by block identity, slow drift, practice effects, or the marginal distribution of block-level RT values.

We repeated the same analysis in the independent online cohort collected using shorter blocks. As shown in **Fig. S3C–D**, observed Rhythm Index values again exceeded the within-subject shuffle-null medians under both destroy-ABAB (one-sided Wilcoxon signed-rank test, p = 5.79 × 10−11) and preserve-ABAB (one-sided Wilcoxon signed-rank test, p = 2.10 × 10−6) controls. These findings indicate that block-locked rhythmicity is a robust feature of BDIAT performance across task formats and data-collection settings.

Together, these shuffle analyses support the conclusion that the rhythmic structure identified in the main text reflects behavioral entrainment to the alternating cognitive demands of the task rather than generic autocorrelation or chance ordering of block-level RT values. The stronger rhythmic entrainment observed in SI− participants may reflect more stable encoding of task context across blocks.

A

D

**Figure S3. Participant-level Comparison of Observed Rhythm Index Relative to Shuffle-derived Null Distributions. (A)** Distribution of participant-level differences between the observed Rhythm Index and the modal value obtained from within-subject shuffles that destroy the ABAB block order in the primary cohort (n=77). Positive values indicate that the observed rhythmic structure exceeds that expected under block-order–randomized nulls. The distribution is shifted above zero (one-sided Wilcoxon signed-rank test$, p = 5.87 \times{10}^{-12}$), indicating that the rhythmic pattern depends on the task-defined alternation of block types. **(B)** Distribution of participant-level differences between the observed Rhythm Index and the modal value obtained from shuffles that preserve the ABAB structure while randomizing the order of blocks within each category in the primary cohort. Although the ABAB structure is maintained, differences remain significantly greater than zero (one-sided Wilcoxon signed-rank test$, p = 0.0068$), indicating that the observed rhythmic pattern is not fully explained by block identity alone. **(C)** Distribution of participant-level differences between the observed Rhythm Index and the modal value obtained from destroy-ABAB shuffles in the online cohort (n=83). As in the primary cohort, values are significantly greater than zero (one-sided Wilcoxon signed-rank test,$p = 5.79 \times10⁻¹¹$), demonstrating robust task-locked rhythmicity under shorter block lengths. **(D)** Distribution of participant-level differences between the observed Rhythm Index and the modal value obtained from preserve-ABAB shuffles in the online cohort. Differences remain significantly greater than zero (one-sided Wilcoxon signed-rank test, $p = 2.10 \times{10}^{-6}$), indicating that the structured temporal dynamics extend beyond simple ABAB block identity and generalize across task formats.

B

C

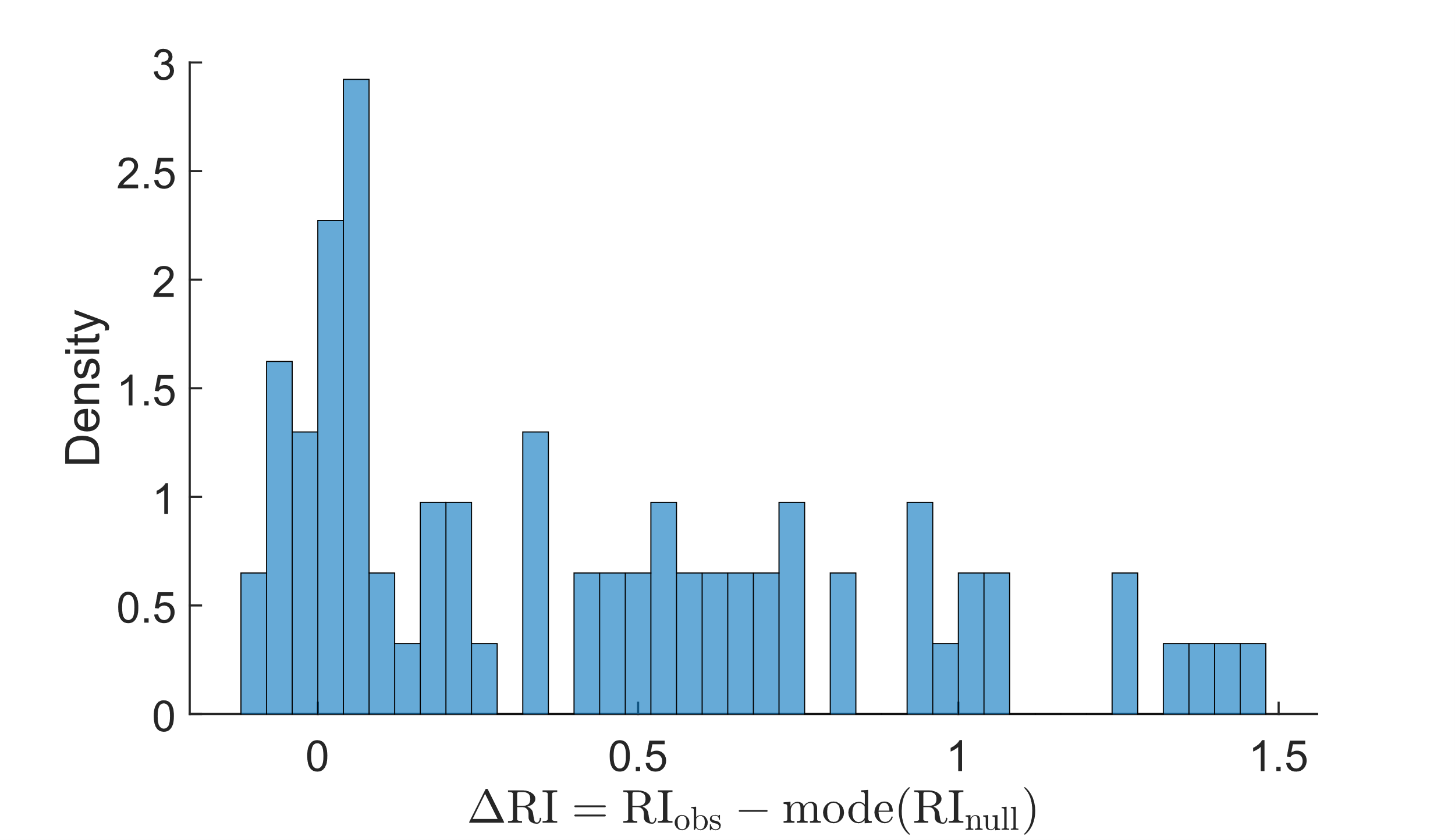

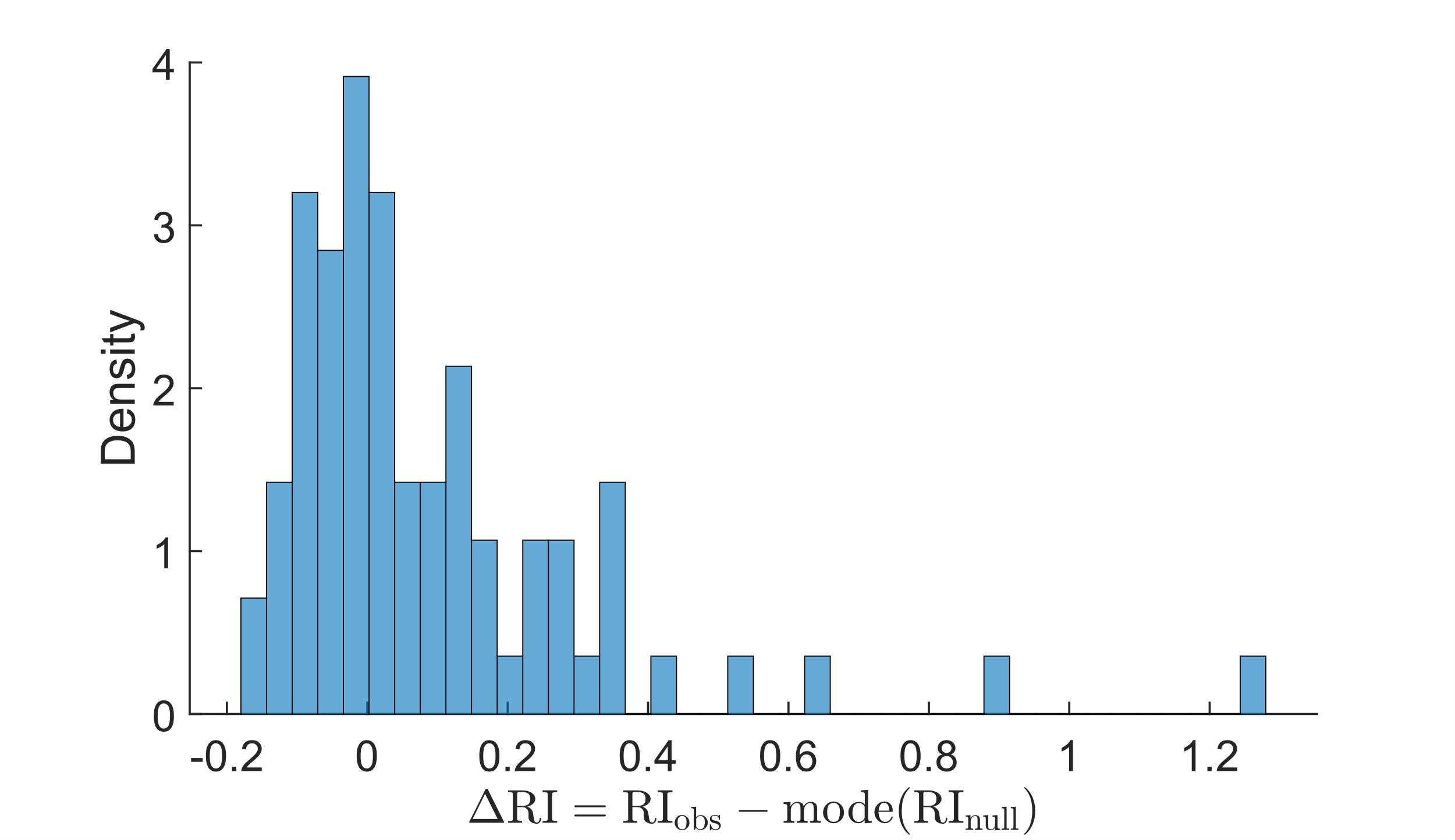

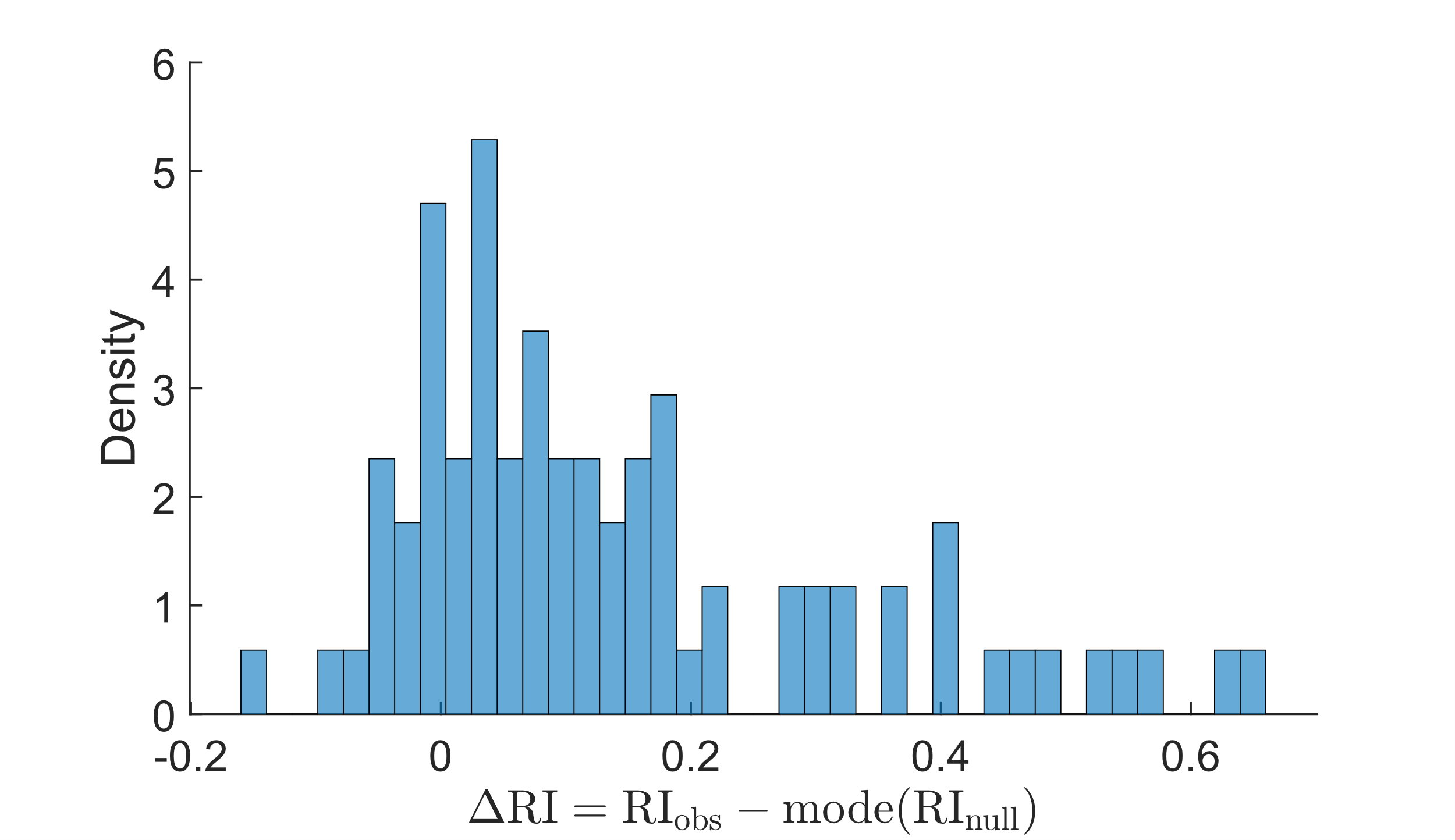

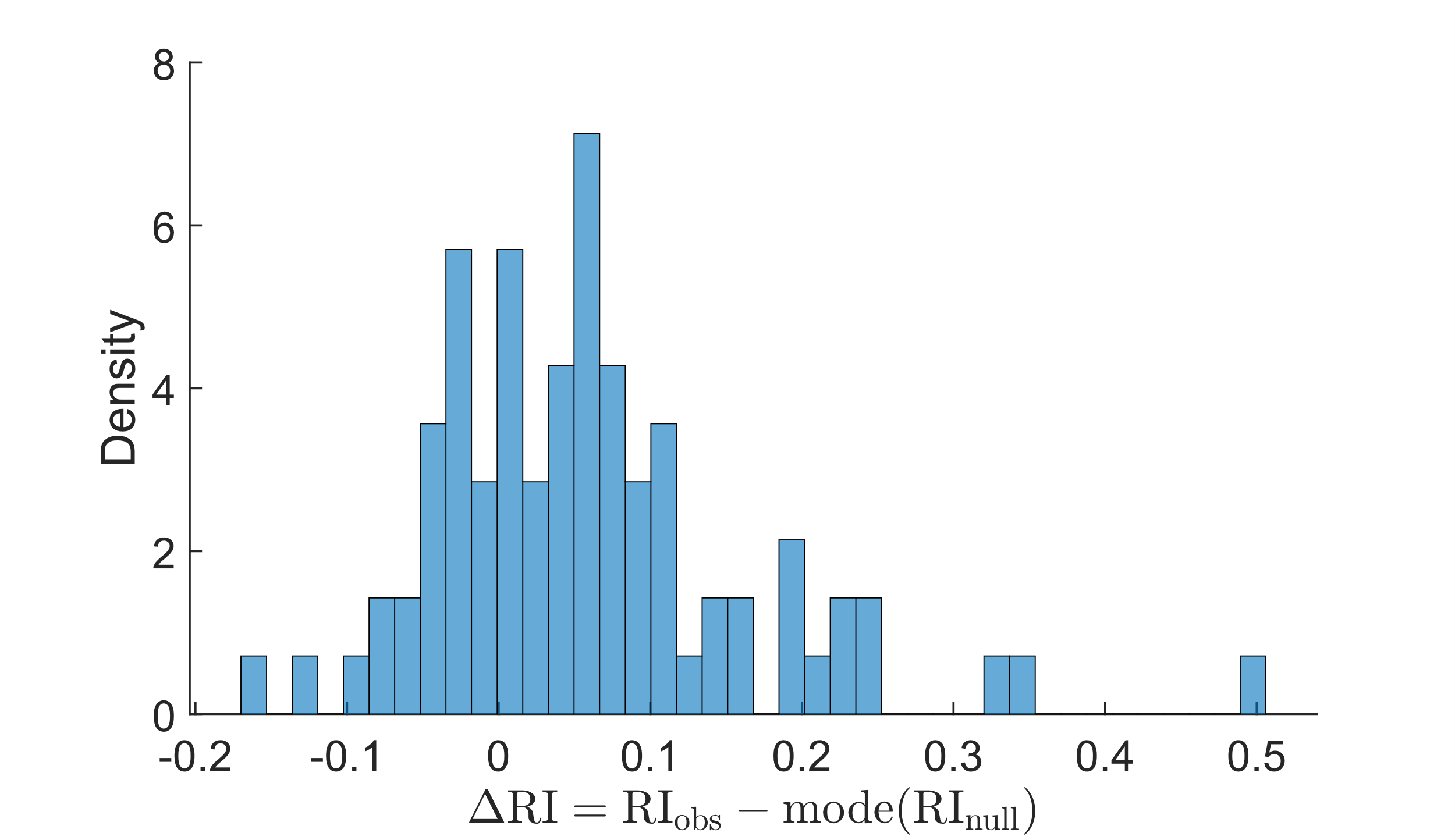

**Section 11. Group Comparisons Across Trials.** For visualization and exploratory assessment, trial-wise two-sample t-tests were conducted to compare mean RTs between SI+ and SI− participants at each of the 20 trial indices within a block. Because these comparisons involved repeated testing across ordered trial positions, P-values were adjusted using the Benjamini–Hochberg FDR procedure. Statistical significance was assessed using FDR-adjusted P-values.

**Section 12. Mixed-Effects Modeling.** To evaluate group differences across the full temporal profile of RT dynamics, we employed a linear mixed-effects model of the form:

$x_{ij} = \beta₀ + \beta₁\cdot Group_{i} + \beta₂\cdot Trial_{j} + \beta₃\cdot(Group_{i} \times Trial_{j}) + u_{i} + \varepsilon_{ij}$*,*	(7)

where $i$ indexes participants and $j$ indexes trial position within a block. The dependent variable, $x_{ij}$, denotes the latent RT-related state for participant $i$ at trial $j$, inferred from the log-transformed RT observations using the preprocessing and state-space modeling pipeline described above. Accordingly, $x_{ij}$ represents the estimated underlying state relative to the participant-specific baseline rather than the observed RT itself. Fixed effects include Group (SI+ and SI−), Trial (trial index), and their interaction. The term $u_{i}$ represents participant-level random intercepts, assumed to be normally distributed with mean zero, and $\epsilon_{ij}$ denotes residual error. To allow block-specific effects while avoiding higher-order interactions across block types, we fit the model separately for the “Death + Me” and “Life + Me” blocks.

Significance of fixed effects was assessed using likelihood-ratio tests comparing nested models(15). Planned contrasts were conducted at trials 1–6 to probe early post-switch dynamics, focusing on the initial adaptation period following the block transition; trial 1 was excluded because it primarily reflects the immediate switch cost. To account for multiple comparisons within this predefined subset, P-values were adjusted using the Benjamini–Hochberg FDR procedure(16).

**Section 13. Uncentered Covariance Matrix Calculation and PCA in Trial.** To characterize latent structure in the RT trajectories, we applied uncentered PCA to participant-specific RT time series. For each participant, RT trajectories from the 9 “Death + Me” blocks were arranged into a matrix $\mathbf{Z}\in\mathbb{R}^{9\times20}$, where each row corresponds to one block-level realization and each column corresponds to one trial position within a block. Because uncentered PCA depends on the scale of the input trajectories, we applied this procedure not only to the raw RT-scale representation $\mathbf{Z}$, but also to elementwise transformed representations of the same trajectories. In addition to the log-transformed representation $\mathbf{X}=log\mathbf{Z}$, defined in the Preprocessing and Normalization section, we considered a family of inverse-power transformations, $\mathbf{Y}^{\left( \alpha\right)}=\mathbf{Z}^{-\alpha},$applied elementwise, where $\alpha>0$ is a tuning parameter and $\alpha=1$ corresponds to the reciprocal transform. These alternative representations were introduced to assess whether the latent structure identified by uncentered PCA was robust to nonlinear rescaling of RTs.

The inverse-power family was motivated by the observation that normalized RT-scale values typically fluctuate around 1. Transformations of the form $z\mapsto z^{-\alpha}$ therefore preserve values near this operating range while compressing larger RT values more strongly than smaller ones. As a result, large single-trial excursions contribute less strongly to the second-moment structure underlying uncentered PCA, while the overall temporal organization of the trajectories is preserved. For notational convenience, let $\mathbf{W}\in\mathbb{R}^{9\times20}$ denote a participant-specific trajectory matrix under any representation considered here, so that $W$may correspond to $\mathbf{Z}$, $\mathbf{X}$, or $\mathbf{Y}^{\left( \alpha\right)}$.

For each participant and each representation $\mathbf{W}$, uncentered PCA was applied separately to the corresponding $9\times20$ trajectory matrix. Let $\boldsymbol{w}_{j}=\mathbf{W}_{\left( j,: \right)}^{\top}\in\mathbb{R}^{20}$denote the column-vector form of the $j$th row of $\mathbf{W}$. In uncentered PCA, principal components are aligned with directions of maximal second-moment variation and therefore reflect contributions from both the mean structure and the covariance of the trajectories. The first principal component (PC1), denoted $\bar{\boldsymbol{v}}$, was obtained by solving

$\bar{\boldsymbol{v}}=arg\underset{\parallel\boldsymbol{v}\parallel=1}{max}\boldsymbol{v}^{\boldsymbol{\top}}\mathbf{S}\boldsymbol{v}=arg\underset{\parallel\boldsymbol{v}\parallel=1}{max}\left( \boldsymbol{v}^{\top}\tilde{\boldsymbol{\Sigma}}\boldsymbol{v} + \boldsymbol{v}^{\boldsymbol{\top}}\tilde{\boldsymbol{\mu}}{\tilde{\boldsymbol{\mu}}}^{\boldsymbol{\top}}\boldsymbol{v} \right)$*,*	(8)

where $\mathbf{S}$, $\tilde{\boldsymbol{\mu}}$, and $\tilde{\boldsymbol{\Sigma}}$ are defined as

$\mathbf{S}=\tilde{\boldsymbol{\Sigma}}+\tilde{\boldsymbol{\mu}}{\tilde{\boldsymbol{\mu}}}^{\boldsymbol{\top}}=\frac{1}{9}\sum_{j=1}^{9} \boldsymbol{w}_{j}\boldsymbol{w}_{j}^{\top}$*,*	(9a)

$\tilde{\boldsymbol{\mu}}=\frac{1}{9}\sum_{j=1}^{9} \boldsymbol{w}_{j}$*,*	(9b)

$\tilde{\boldsymbol{\Sigma}}=\frac{1}{9}\sum_{j=1}^{9} (\boldsymbol{w}_{j}-\tilde{\boldsymbol{\mu}})(\boldsymbol{w}_{j}-\tilde{\boldsymbol{\mu}})^{\top}.$	(9c)

Accordingly, the quantity ${\bar{\boldsymbol{v}}}^{\boldsymbol{\top}}\mathbf{S}\bar{\boldsymbol{v}}$represents the total second-moment variation explained along PC1 and decomposes into a covariance contribution, ${\bar{\boldsymbol{v}}}^{\boldsymbol{\top}}\tilde{\boldsymbol{\Sigma}}\bar{\boldsymbol{v}}$, and a mean contribution, ${\bar{\boldsymbol{v}}}^{\boldsymbol{\top}}\tilde{\boldsymbol{\mu}}{\tilde{\boldsymbol{\mu}}}^{\boldsymbol{\top}}\bar{\boldsymbol{v}}$. Applying this procedure separately to each participant yielded participant-specific principal component trajectories, with PC1 representing the dominant latent mode of variation across the 20 trial positions.

To assess whether task-related structure extended beyond the dominant latent dimension, we also examined higher-order components, particularly the second principal component (PC2), using the same participant-wise procedure. The resulting principal component trajectories from each representation were then carried forward to the inferential analyses described in the preceding section. Further analyses of the dependence of the uncentered PCA structure on RT transformations, including the decomposition of mean and covariance contributions and associated inferential results, are provided in Supplementary Information, **Section 14**.

**Section 14. Additional Analysis of Uncentered PCA Across RT Transformations.** Using the participant-wise uncentered PCA procedure described in **Section 13** above, we further examined how the latent structure of RT trajectories depended on the scale on which those trajectories were represented. The analysis focused on RT trajectories from the “Death + Me” blocks and compared the raw RT-scale representation $\mathbf{Z}$, the log-transformed representation $\mathbf{X}=\log\mathbf{Z}$, and the inverse-power family $\mathbf{Y}^{\left( \alpha\right)}=\mathbf{Z}^{-\alpha}$.

For the raw RT-scale representation $\mathbf{Z}$, the first principal component (PC1) explained $95.3\pm1.7\%$ of the total second-moment variation, indicating that participant-specific RT trajectories were strongly aligned with a dominant one-dimensional structure. Of the variation captured by PC1, $98.5\pm1.1\%$ was attributable to the mean contribution, suggesting that PC1 provided a stable latent representation of the participant-specific RT trajectories. Trial-wise two-sample $t$-tests applied to the PC1 trajectories did not reveal evidence for SI-state encoding after correction for multiple comparisons, with the smallest adjusted $q$-value observed at trial 4 ($q=0.36$). Because these tests treat trials independently and do not account for repeated measurements within participants, we also fit linear mixed-effects models to the PC1 trajectories. These analyses did not reveal significant effects of Group, Trial, or Group $\times$ Trial interaction ($P>0.05$ for all terms).

We next examined whether nonlinear rescaling of RT trajectories altered the latent structure recovered by uncentered PCA. For the log-transformed representation $X=\log\mathbf{Z}$, the dominance of PC1 was substantially reduced: the explained variation dropped to $35.2\pm9.4\%$, and only $22.9\pm22.8\%$ of the variation captured by PC1 was attributable to the mean contribution. This shift indicates that the log transform emphasizes covariance-driven structure at the expense of the strong mean-aligned component present in the raw RT-scale representation. Trial-wise group comparisons of the resulting PC1 trajectories again did not survive correction for multiple comparisons, although the smallest adjusted $q$-value remained at trial 4.

We then examined the inverse-power family $\mathbf{Y}^{\left( \alpha\right)}=\mathbf{Z}^{-\alpha}$, which was introduced in the main text to assess the robustness of the latent representation to nonlinear rescaling. For the reciprocal transform ($\alpha=1$), PC1 explained $96.7\pm1.2\%$ of total variation, slightly exceeding the value obtained for the raw RT-scale representation, and the mean contribution within PC1 increased to $99.2\pm0.6\%$. Under this transformation, trial-wise analysis of PC1 revealed the strongest group separation at trial 4 ($q=0.05$). Across $\alpha$values in the range $0.5\leq\alpha\leq2$, the strongest evidence for group-level separation was observed for values near $\alpha=1$.

A plausible explanation for this effect follows from the range of normalized RT-scale values, which typically fluctuates around 1. Transformations of the form $z\mapsto z^{-\alpha}$ preserve values near this operating range while compressing larger RT values more strongly than smaller ones. As a result, unusually large single-trial excursions contribute less strongly to the second-moment structure underlying uncentered PCA, while the dominant mean structure is preserved. Under a first-order Taylor approximation of the map $g(z)=z^{-\alpha}$ around the mean trajectory, the transformed covariance is approximately a locally rescaled version of the original covariance. In practice, this compression appears to reduce the influence of single-trial noise on the PCA objective and thereby sharpen the group-related structure that is most evident near trial 4.

To determine whether SI-related information extended beyond the dominant latent dimension, we also examined higher-order components, particularly the second principal component (PC2). In contrast to PC1, PC2 did not show reliable early-trial separation, and trial-wise differences observed at later trials did not remain significant after correction for multiple comparisons. PC2 trajectories and associated trial-wise $P$- and $q$-values are shown in **Fig. S4**.

**Figure S4. Trial-by-trial Group Comparisons of First Principal Component (PC1) and Second Principal Component (PC2) Loadings**. **(A)** shows the false discovery rate (FDR)-adjusted q-values for PC1 loadings derived from “Death + Me,” with strong encoding evidence in trial 4 where the q-value reaches statistical significance (q = 0.05). Toward the end of the block, there is evidence of encoding, but it does not reach meaningful statistical significance after correction. **(B)** FDR-adjusted q-values for PC2 loadings derived from “Death + Me” blocks. In contrast to PC1, no trial exhibits statistically significant group differences after multiple-comparison correction. Although some trials display effects, these do not remain statistically significant following FDR adjustment, indicating the absence of consistent encoding evidence in PC2 across the block. A horizontal dashed line indicates the nominal significance threshold (α = 0.05).

B

A

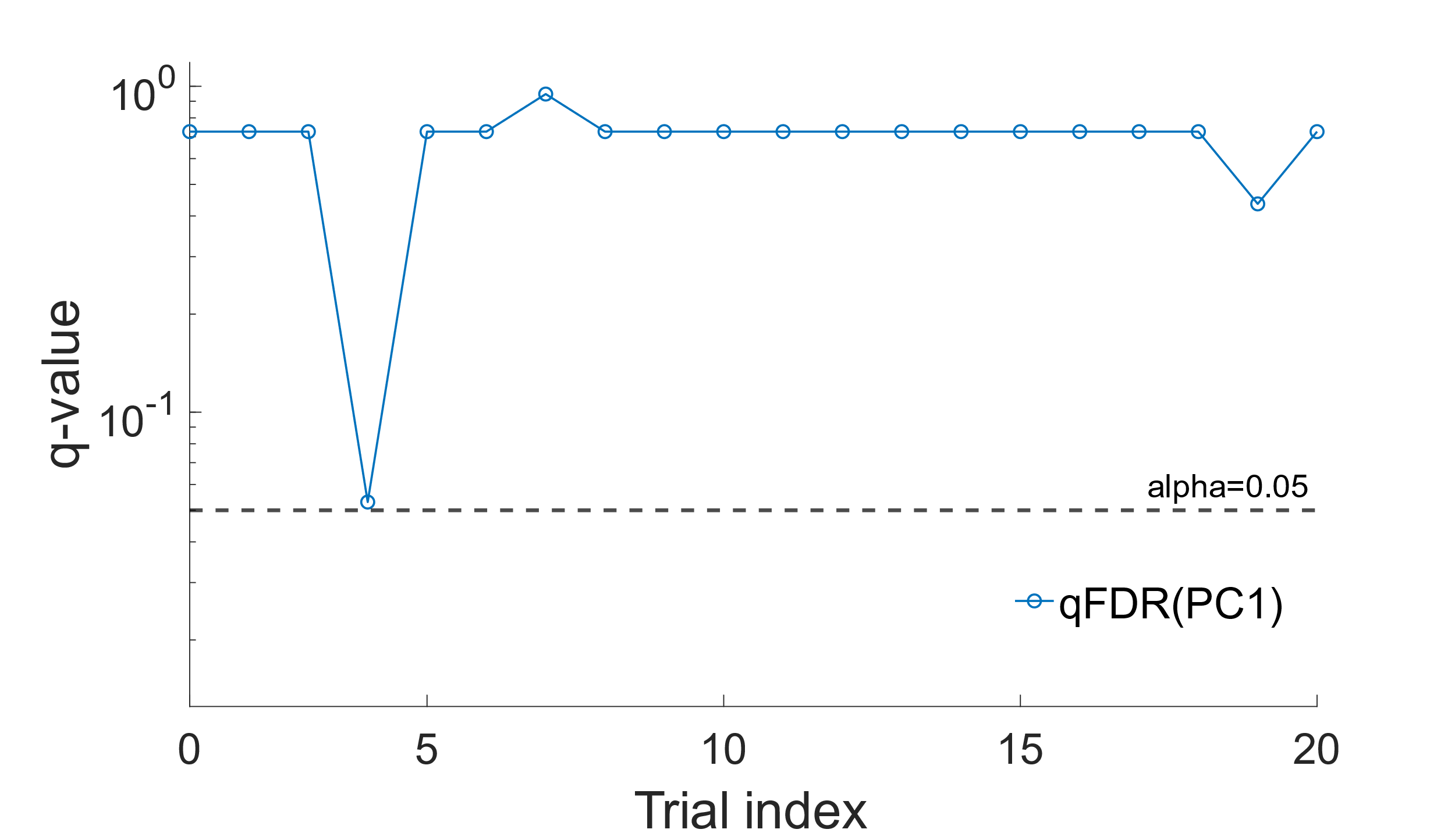

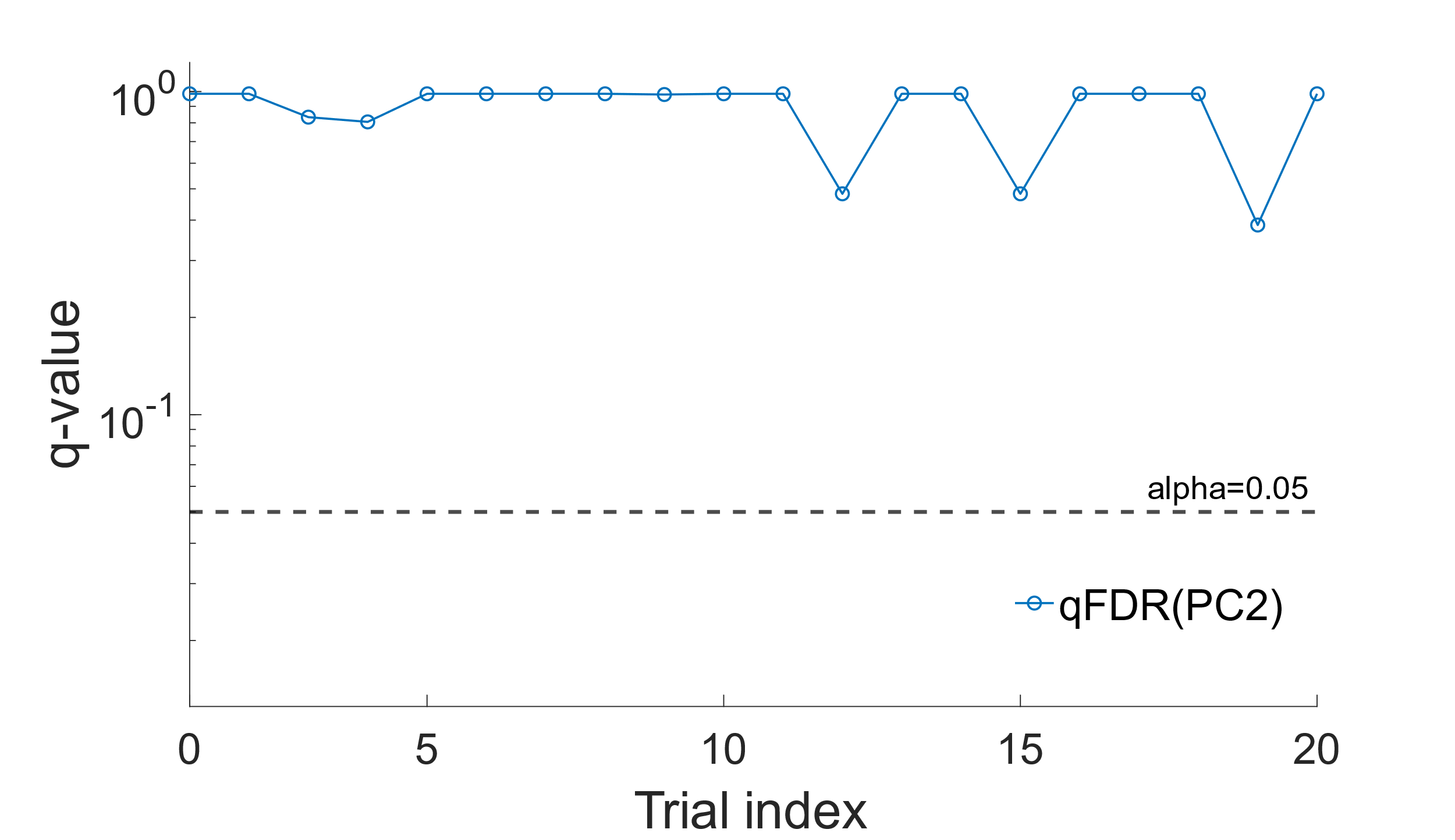

**Section 15. Assessing Uniformity of Stimulus Across Trials and Blocks.** In the Results section, we showed that SI status is mainly reflected in early trials right after the block switch. Specifically, this encoding is most evident at trial 4 in “Death + Me” blocks, with partial evidence of encoding in the adjacent trials, namely trials 3, 5, and 6. The concentration of temporal encoding at these specific trials raises the concern that the effect might arise spuriously because particular stimuli happen to be disproportionately presented at trial 4 or its adjacent trial indices. To exclude this possibility, we examined whether particular stimulus words were preferentially presented at these trial indices within “Death + Me” blocks.

In particular, we tested whether the composition of stimulus words differed systematically across trial indices. For a given trial position, we first formed the observed count vector by summing, across all participants, how many times each stimulus word was presented at that position, where the categories correspond to the individual words themselves (e.g., “self,” “mine,” “live,” and “die”). Let $n_{t}$ denote the total number of word presentations at that trial. We then constructed a reference distribution by pooling counts of the same words across all other trial positions within the block and converting those totals into probabilities. Under the null hypothesis that stimulus allocation is homogeneous across positions, the counts at the target trial should be well described as draws from a multinomial distribution with sample size $n_{t}$ and category probabilities given by this pooled distribution.

We quantified the discrepancy between the observed counts and the expected counts ($n_{t}$ multiplied by the pooled probabilities) using the Pearson chi-square goodness-of-fit statistic(17). Because some categories have relatively small expected frequencies, we assessed significance using Monte Carlo sampling. Specifically, we generated many synthetic count vectors for trial $t$ by repeatedly drawing multinomial samples with the same $n_{t}$ and pooled probabilities. For each synthetic sample, we recomputed the chi-square statistic, thereby obtaining the distribution of mismatch values expected purely from random variation in word assignment. The Monte Carlo p-value corresponds to the proportion of simulated statistics that were at least as large as the empirical one. Across all trial positions, including those showing the strongest temporal encoding in the main analyses, trials 1, 3, 4, 5, and 6, we observed no significant departures from the pooled distribution.

**Fig. S5A** illustrates the relative frequency of each stimulus across trial index, which shows to be reasonably homogeneous allocation throughout the whole block. **Fig. S5B** shows the null distribution and the observed statistic for trial index 4. The observed value lies well within the range expected under the null hypothesis; thus, it indicates that stimulus word presentation does not account for the temporal effects observed in our analysis.

**Figure S5. Control Analysis for Stimulus-specific Effects at Early Trial Indices within “Death + Me” Blocks. (A)** Heatmap showing the probability of each stimulus word as a function of trial index, computed across all participants. Colors indicate probability density. The distribution of stimulus words is reasonably uniform across trial indices, with no clear preference or lack of specific words at early trials (vertical line denotes trial 4). **(B)** Monte-Carlo evaluation of the Pearson chi-square goodness-of-fit statistic tests whether the distribution of stimulus words at trial index 4 differs from the distribution pooled across all other trial indices. The histogram shows the null distribution of chi-square statistics obtained by repeatedly generating synthetic trial-4 count vectors (summed across participants) from a multinomial distribution defined by the pooled probabilities of the remaining trials and matched in total sample size. The dashed line marks the observed statistic computed from the actual trial-4 data. The observed value falls well within the null distribution, indicating that the stimulus composition at trial 4 is statistically indistinguishable from that of the remaining trials.

B

A

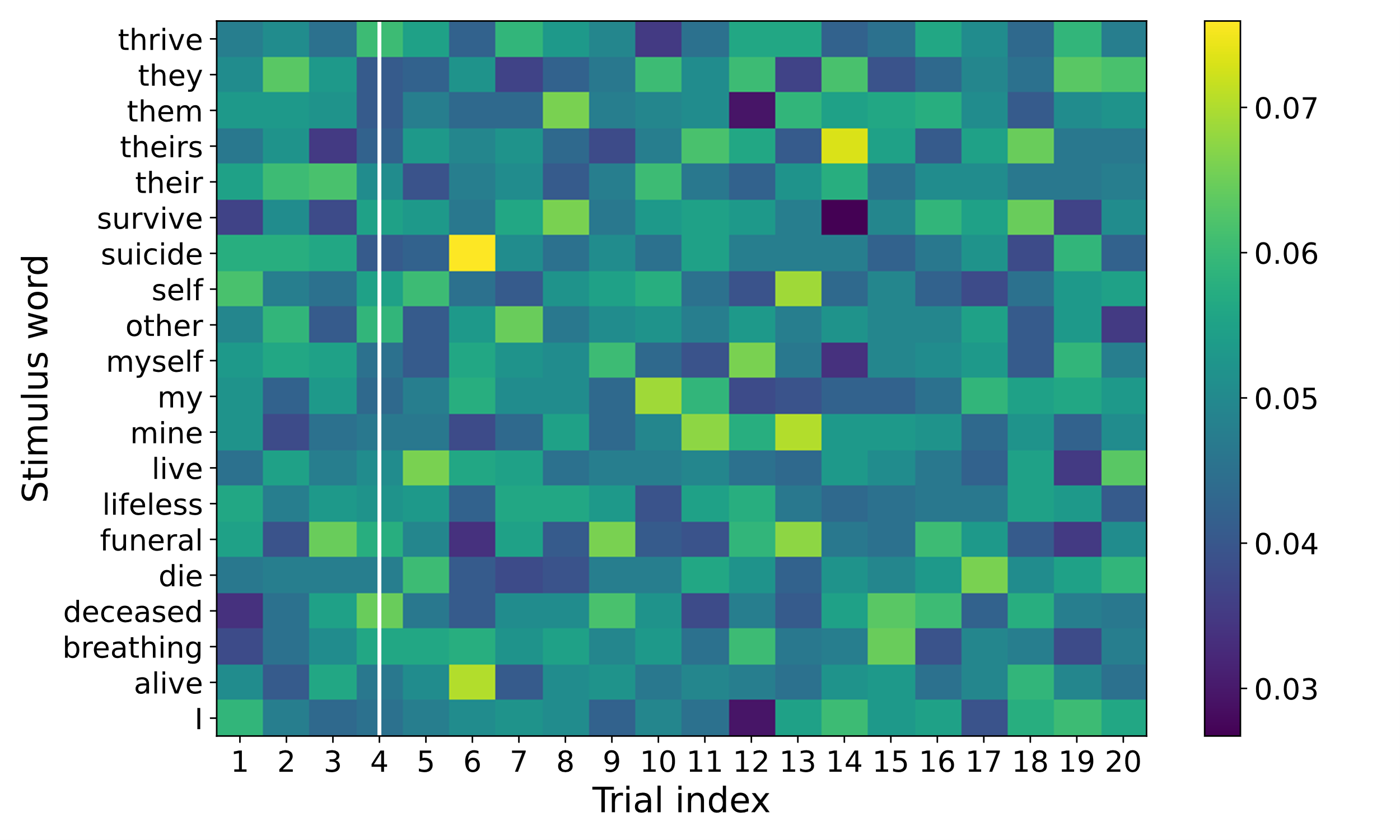

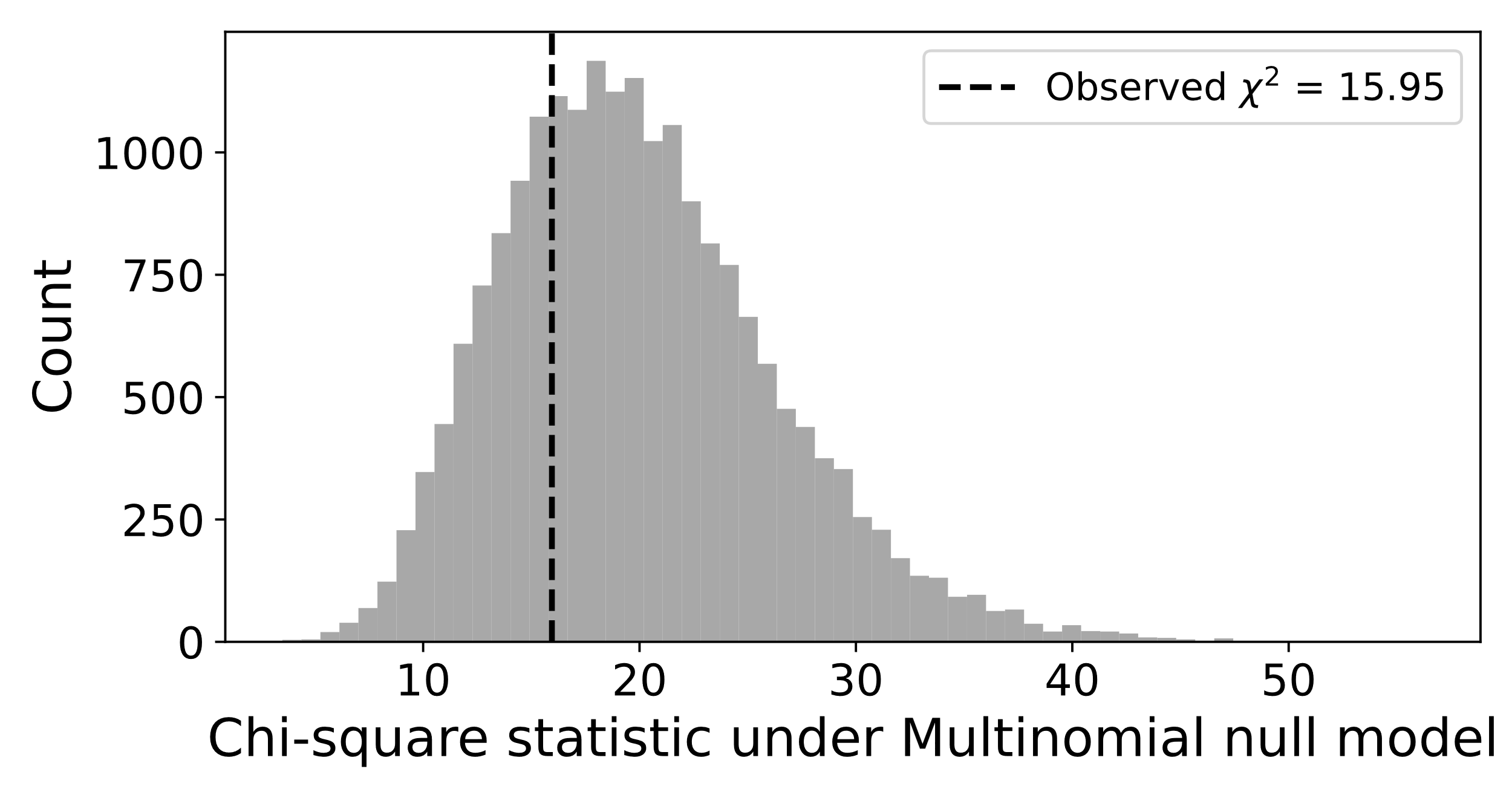

**Section 16. Bilinear Logistic Regression Classifier.** To model how temporal structure in RT data relates to SI state, we developed a bilinear logistic regression framework that jointly captures block-level organization and within-block temporal dynamics. The model is motivated by our preliminary analyses showing that SI-related differences are expressed both across alternating task blocks and within fine-grained trial-level RT structure. Accordingly, the model decomposes each participant’s RT time series into separable block-level and trial-level components, enabling interpretable characterization of spatio-temporal behavioral patterns. The classifier is built using “Death + Me” blocks because prior analyses indicated that SI-related temporal structure and autocorrelation effects were strongest in these blocks, making them the most informative segment for decoding SI state. This formulation directly operationalizes temporal dependencies identified in earlier analyses.

For each participant $i$, RT time series data from the “Death + Me” blocks were represented as a matrix $\mathbf{Z}_{i}\in\mathbb{R}^{9\times20}$, and SI state was encoded as a binary outcome variable $y_{i}\in\{0,1\}$, with $y_{i}=0$denoting SI− and $y_{i}=1$ denoting SI+. The log-odds of SI+ are then defined as:

$\log\left( \frac{p_{i}}{1-p_{i}} \right)=b_{0}+\sum_{j=1}^{J} \mathbf{b}_{j}^{T}\mathbf{Z}_{i}^{\boldsymbol{-}\alpha}\mathbf{v}_{j},$	(10)

where $b_{0}$ denotes the intercept, $\mathbf{b}_{j}\in\mathbb{R}^{9\times1}$ are block-level weighting vectors, $\mathbf{v}_{j}\in\mathbb{R}^{20\times1}$ are trial-level weight vectors and $\alpha$ is set close to 1. The integer J denotes the number of latent spatiotemporal components. Here, $p_{i}$ denotes the modeled probability of belonging to the SI+ under the bilinear logistic regression model defined above.

The classifier parameters $\Theta= \{b_{0}, \mathbf{b}_{\left\{ 1:J \right\}}, \mathbf{v}_{\left\{ 1:J \right\}}\}$were learned by maximizing the weighted log-likelihood function:

$\hat{\Theta}=\underset{\Theta}{\mathrm{argmax}} \sum_{i=1}^{N} \omega_{i}\left[ y_{i}log(p_{i})+(1-y_{i}) log(1-p_{i}) \right],$	(11a)

$subject to \left\{ \begin{aligned} \mathbf{b}_{j}^{T}\mathbf{b}_{j}=1 j=1, \ldots, J, \\ \mathbf{b}_{j}^{T}\mathbf{b}_{k}=0 j\neq k , J\geq2 \\ \sum_{j=1}^{J} \left\| \mathbf{v}_{j} \right\|_{1}\leq C. \end{aligned} \right.,$	(11b)

The term $\omega_{i}$ represents class-specific sample weights applied to the log-likelihood to correct for group-size imbalance. The vectors $\mathbf{b}_{j}$ are the block-level loading vectors, $\mathbf{v}_{j}$ are the trial-level weight vectors, and $b_{0}$is the intercept term, estimated jointly with remaining parameters. The parameter $C$ sets the bound on $\left\| \mathbf{v}_{j} \right\|_{1}$and thus controls sparsity in temporal weights. This regularization reflects our empirical observation that SI-related effects are localized to specific trial indices, and it also mitigates overfitting given the small sample size of our dataset. Block-level weight vectors $\mathbf{b}_{j}$ were constrained to have unit norm to prevent scale indeterminacy between block and trial contributions, and for models with $J\geq2$, were additionally constrained to be orthogonal to ensure distinct component representations.

Parameter estimation proceeded via an alternating two-step optimization procedure. First, holding $\mathbf{b}_{j}$ fixed, we constructed standardized features and estimated $\mathbf{v}_{j}$and $b_{0}$ using $\mathcal{l}_{1}$-regularized logistic regression (GLM-LASSO)(18), with the regularization strength selected by K-fold cross-validation within the training data using one-standard-error rule(19) by default. Specifically, a regularization path corresponding to different levels of $\mathcal{l}_{1}-$ regularization strength was computed within each training fold, and the optimal regularization level was selected based on cross-validated deviance. This subproblem is convex in $\mathbf{v}_{j}$ for fixed $\mathbf{b}_{j}$ and therefore provides a unique optimum at each alternation. Second, holding $\mathbf{v}_{j}$ and $b_{0}$ fixed, we updated each $\mathbf{b}_{j}$using gradient-ascent steps on the weighted likelihood function, followed by renormalization and (when $J\geq2$ ) Gram-Schmidt orthogonalization. A fixed number of outer alternations and gradient steps per alternation were performed using a constant step size, without adaptive stopping criteria. Because the overall bilinear objective is non-convex, the alternating procedure converges to a local optimum.

The number of bilinear components $J$ was treated as a model hyperparameter and evaluated using cross-validation. Candidate values of $J$ were compared based on LOOCV performance, and the value yielding the highest balanced accuracy was selected. Predictive performance was assessed using LOOCV, in which each participant was held out in turn, the model was trained on the remaining $N-1$ participants, and SI state (SI+ vs SI-) was predicted for the held-out participant. Classification metrics, including balanced accuracy, sensitivity, specificity, and AUC, were computed by aggregating predictions across all folds. This procedure provides an estimate of generalization to unseen participants while maintaining a compact and interpretable parameterization.

To assess whether classification performance exceeded chance, we conducted a nonparametric label-shuffling permutation analysis with $1,000$ iterations. For the primary analysis under true labels, optimization parameters (e.g., number of gradient steps and step size) were fixed a priori. In contrast, for each permuted dataset, we evaluated a predefined grid of optimization settings and retained the best-performing configuration within that grid, i.e. we specifically biased this analysis towards the null finding. For each shuffle, SI labels were randomly reassigned across participants and the entire LOOCV training and evaluation pipeline, including regularization selection within each fold, was repeated from scratch. Balanced accuracy and AUC were computed for each shuffled run to generate empirical null distributions that reflect both model fitting and hyperparameter selection. Empirical P-values were calculated as the proportion of permuted runs achieving performance equal to or greater than that observed under the true labels.

**Section 17. Comparative Analysis with Sequence-based Classifiers.** To assess whether alternative classifier models can predict SI with the same or better performance than our proposed pipeline, we conducted a comparative analysis. For the comparison, we used a set of state-of-the-art classifier models ranging from simpler models including logistic regression, support vector machine (SVM)(15) and multilayer perceptron (MLP). For more complex classifier models, we picked long short-term memory (LSTM)(20), transformers(21), and large language model (LLM)–based classifiers(22). It is worth noting that the effectiveness of these models is strongly dependent on the size and scale of the available data(19,23).

To avoid potential overfitting issues, the comparative analysis incorporated regularization, sparsity-inducing penalties where applicable, and validation-based model selection across all methods. All baseline models were evaluated using participant-level leave-one-out cross-validation (LOOCV; 77 folds), ensuring strict separation between training and test subjects. Performance was quantified using accuracy, balanced accuracy, F1-score, sensitivity, specificity, and area under the receiver operating characteristic curve (AUC). For each classifier family, predefined input representations, including raw trial-level RT, block-averaged RT, and preprocessed RT sequences, were evaluated, with representation selection performed exclusively within training folds to avoid information leakage. No task-specific feature engineering or label-aware preprocessing was used, contributing to a comparatively fair evaluation framework across methods.

**Fig. S6** shows ROC curves of the evaluated classifier models. Linear classifier models generally demonstrated limited discriminative performance relative to the proposed approach, particularly when using block-level summary representations, indicating that low-dimensional aggregate statistics do not adequately capture the temporal structure associated with SI state. Logistic regression (AUC = 0.563), linear SVM models trained with $\mathcal{l}_{1}-$regularization (AUC = 0.617), and the MLP classifier (AUC = 0.586) all showed comparatively limited discrimination, remaining substantially below the performance achieved by the proposed bilinear logistic regression framework.

We further evaluated sequence-aware neural architectures, including LSTM networks, transformer encoders, one-dimensional convolutional neural networks (1D-CNNs)(24) and LLM-based embeddings passed to a logistic regression classifier, adapted to operate on discretized RT tokens. Overall, these sequence-aware architectures achieved moderate discrimination relative to our proposed bilinear logistic regression model. The strongest performance among these models was obtained with the LSTM, which achieved a balanced accuracy of 0.682 and an AUC of 0.712. Although this indicates that recurrent architectures can recover meaningful temporal information from the sequences, their performance remained below that of the proposed approach (balanced accuracy = 0.770; AUC = 0.799). These results suggest the value of incorporating task-informed structure when extracting clinically relevant temporal patterns. Complete performance metrics for all models are summarized in **Table S1**.

**Figure S6. Performance of Baseline Classifier to Classify SI+ vs SI− using RT Time Series.** Models were evaluated using participant-level leave-one-out cross-validation (LOOCV; 77 folds). For each classifier family, predefined input representations, including raw trial-level reaction time (RT), block-averaged RT, and preprocessed RT sequences, were evaluated, with representation selection performed exclusively within training folds to avoid information leakage. Symbols in the legend indicate the best-performing representation selected for each model ($◯$, raw RT; $\square$, block-averaged RT; $\triangle$, preprocessed RT). Linear classifiers operating on block-level summaries exhibited near-chance performance, indicating that low-dimensional aggregate statistics fail to capture the temporal organization associated with SI state. Sequence-based neural architectures demonstrated improved discriminative performance, with the strongest baseline result obtained by the LSTM model (balanced accuracy = 0.682; area under curve (AUC) = 0.712). Nevertheless, all alternative approaches remained below the performance of the proposed bilinear logistic regression framework (balanced accuracy = 0.770; AUC = 0.799). Complete performance metrics for all methods are provided in **Table S1**.

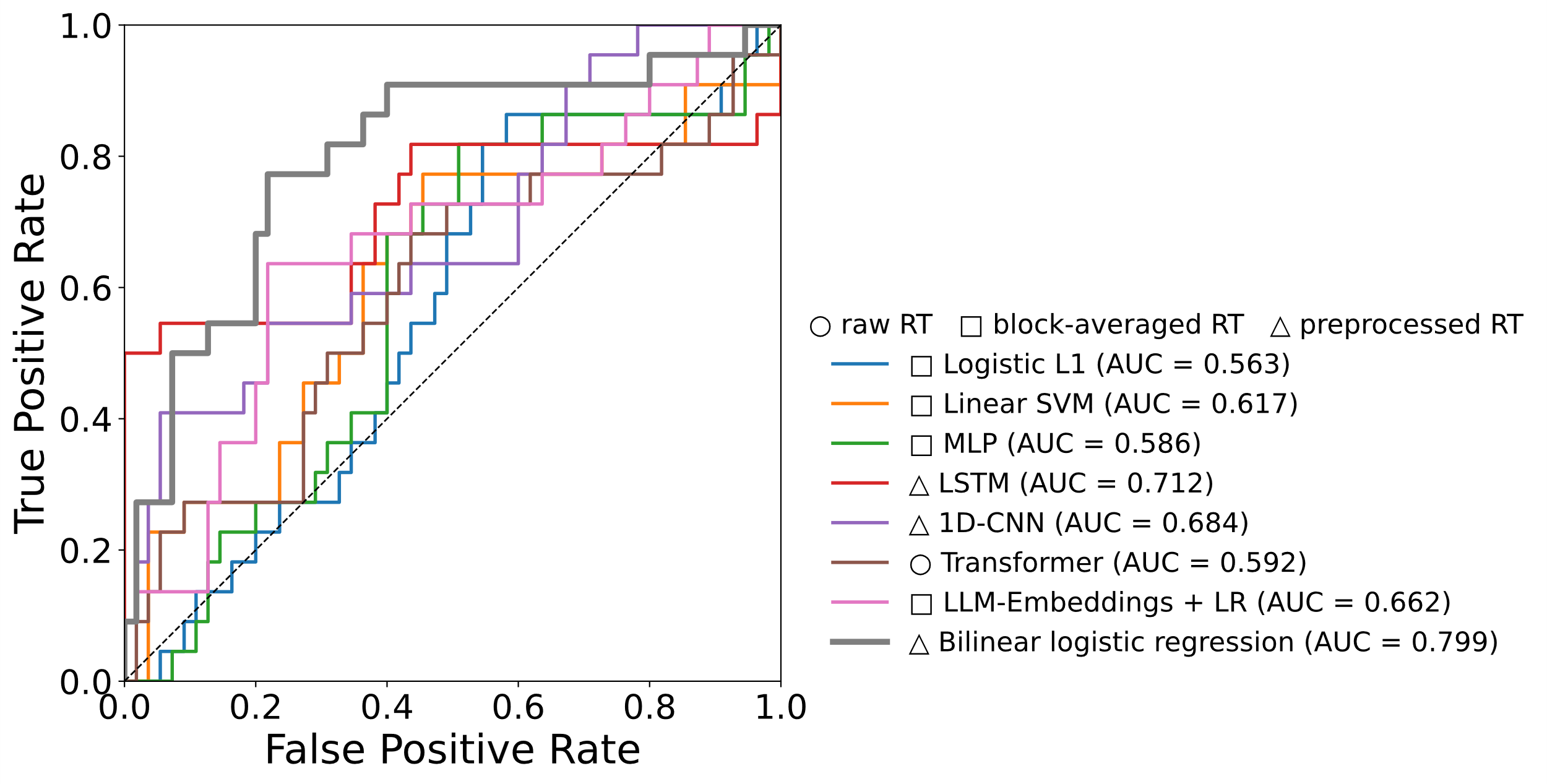

**Table S1. Performance Comparison Between Baseline Classifiers and the Proposed Bilinear Model.** Classification metrics obtained under leave-one-out cross-validation (LOOCV) at the participant level. All models were trained on identical preprocessed reaction time (RT) sequences without access to block structure or task-informed feature engineering. Metrics reported are balanced accuracy, sensitivity, specificity, and area under the receiver operating characteristic curve (AUC). Among the baselines, the long short-term memory (LSTM) achieved the strongest overall performance (balanced accuracy = 0.682; AUC = 0.712), but remained below the performance of the bilinear logistic regression model.

| **Model** | **Balanced Accuracy** | **F1-score** | **Sensitivity** | **Specificity** | **AUC** |
| --- | --- | --- | --- | --- | --- |
| Logistic L1 | 0.532 | 0.386 | 0.500 | 0.564 | 0.563 |
| Linear SVM | 0.550 | 0.419 | 0.591 | 0.509 | 0.617 |
| MLP | 0.677 | 0.548 | **0.773** | 0.582 | 0.586 |
| LSTM | 0.682 | 0.545 | 0.545 | **0.818** | 0.712 |
| 1D-CNN | 0.618 | 0.455 | 0.455 | 0.782 | 0.684 |
| Transformer | 0.523 | 0.318 | 0.318 | 0.727 | 0.592 |
| LLM Embeddings with Logistic Regression Classifier | 0.650 | 0.517 | 0.682 | 0.618 | 0.662 |
| Bilinear logistic regression | **0.770** | **0.667** | **0.773** | 0.782 | **0.799** |

**Section 18. Diagnostic Specificity of Temporal Features.** The primary analyses in the main text focused on predicting near-term SI derived from EMA. Because SI is correlated with psychiatric diagnosis, an important question is whether the temporal features identified by our framework primarily reflect general case–control differences rather than SI-specific dynamics.

To address this possibility, we repeated the full analytical pipeline using diagnostic labels, major depressive disorder (MDD) versus control in place of SI state, SI+ vs SI−. All preprocessing, feature construction, model structure, and cross-validation procedures were kept identical to those used in the main analyses. Because the labels were the only element changed in this step, any differences in results can be attributed to the outcome definition rather than the analytic pipeline.

At the descriptive level, the early-trial divergence observed in **Fig. 3A** of the main text was not present when participants were grouped by diagnosis as shown in **Fig. S7A**. Group-averaged trajectories showed substantial overlap, and no consistent separation emerged at the trial positions previously associated with SI-related modulation.

The SI classifier performance was reduced when diagnostic labels were substituted for SI status. The ROC curve obtained under MDD versus control classification in **Fig. S7C** shows weaker discriminative performance, with an AUC of 0.601 compared to an AUC of 0.799 for SI+ versus SI− classification in **Fig. 4A** of the main text. This reduction in performance indicates that the temporal structure inferred by the prediction model to separate SI+ versus SI− groups does not generalize to discriminating MDD versus control groups.

Together, these results suggest that the behavioral features identified in the main analyses are more tightly linked to short-term fluctuations in SI than to broad diagnostic grouping. These findings further support the conclusion that the model captures state-dependent cognitive dynamics rather than stable trait differences associated with psychiatric diagnosis.

**Figure S7. Evaluations of Reaction Time (RT) Dynamics and Encoding Under Major Depressive Disorder (MDD) vs Control Diagnostic Grouping (A)** Mean RT-scale trajectories within “Death + Me” blocks when participants are grouped by diagnostic status (MDD versus Control). Shaded bands represent bootstrap 95% confidence intervals around the group mean trajectories. In contrast to the SI-based grouping shown in Fig. 3A of the main text, early-trial divergence is not apparent and group averages show substantial overlap across trial positions. **(B)** Trial-wise statistical comparison of the mean RT-scale within “Death + Me” blocks under diagnostic grouping. At each trial position (1–20), we performed two-sample tests with unequal variance and report the Benjamini–Hochberg false discovery rate (FDR)–adjusted q-values. A horizontal dashed line indicates the nominal significance level (α = 0.05). **(C)** Receiver operating characteristic (ROC) curve obtained when the bilinear classification model is trained to predict diagnostic status rather than SI. Discrimination is reduced relative to SI decoding (**Fig. 4A**), indicating that the temporal features captured by the model are not primarily driven by case–control differences.

A

C

B

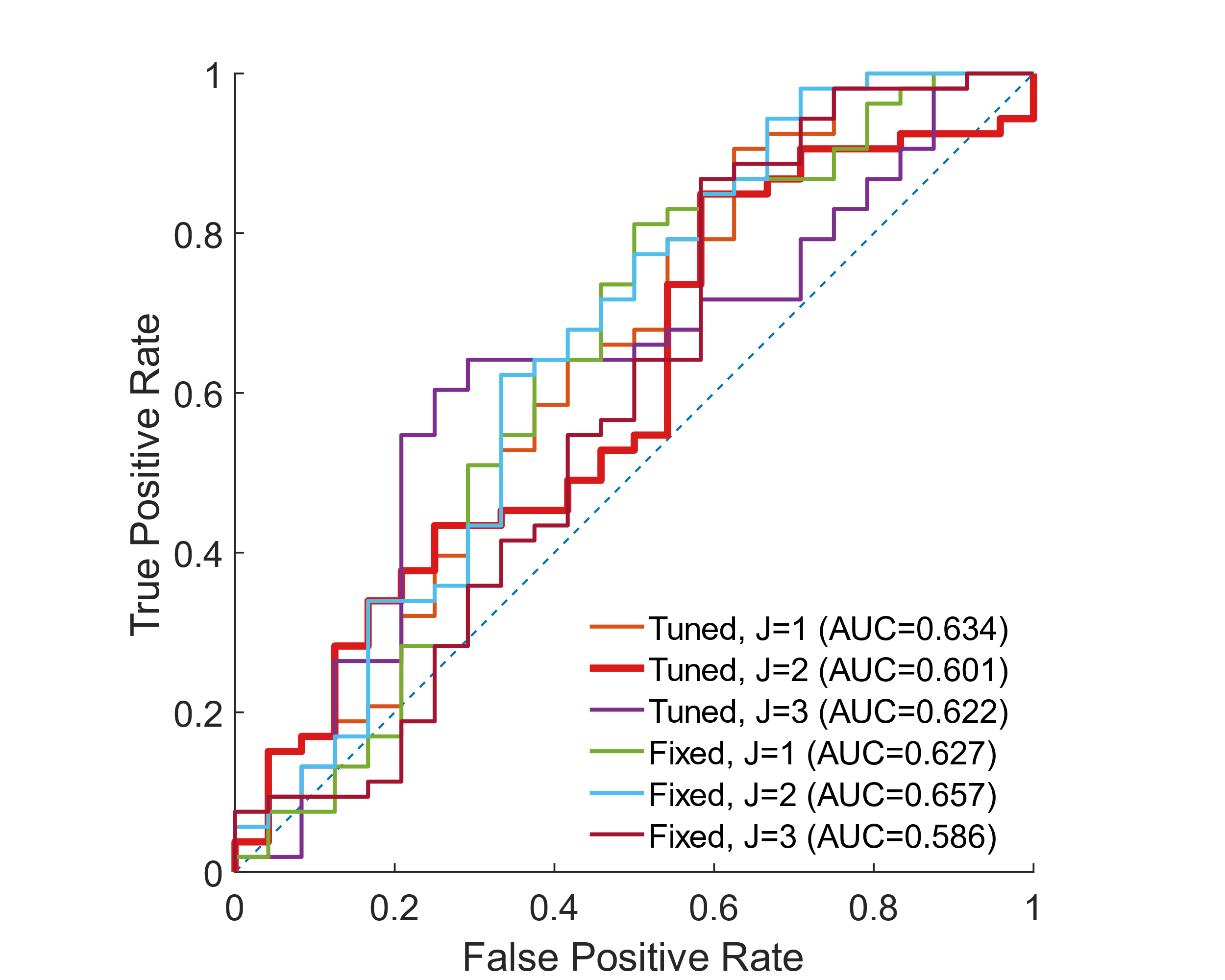

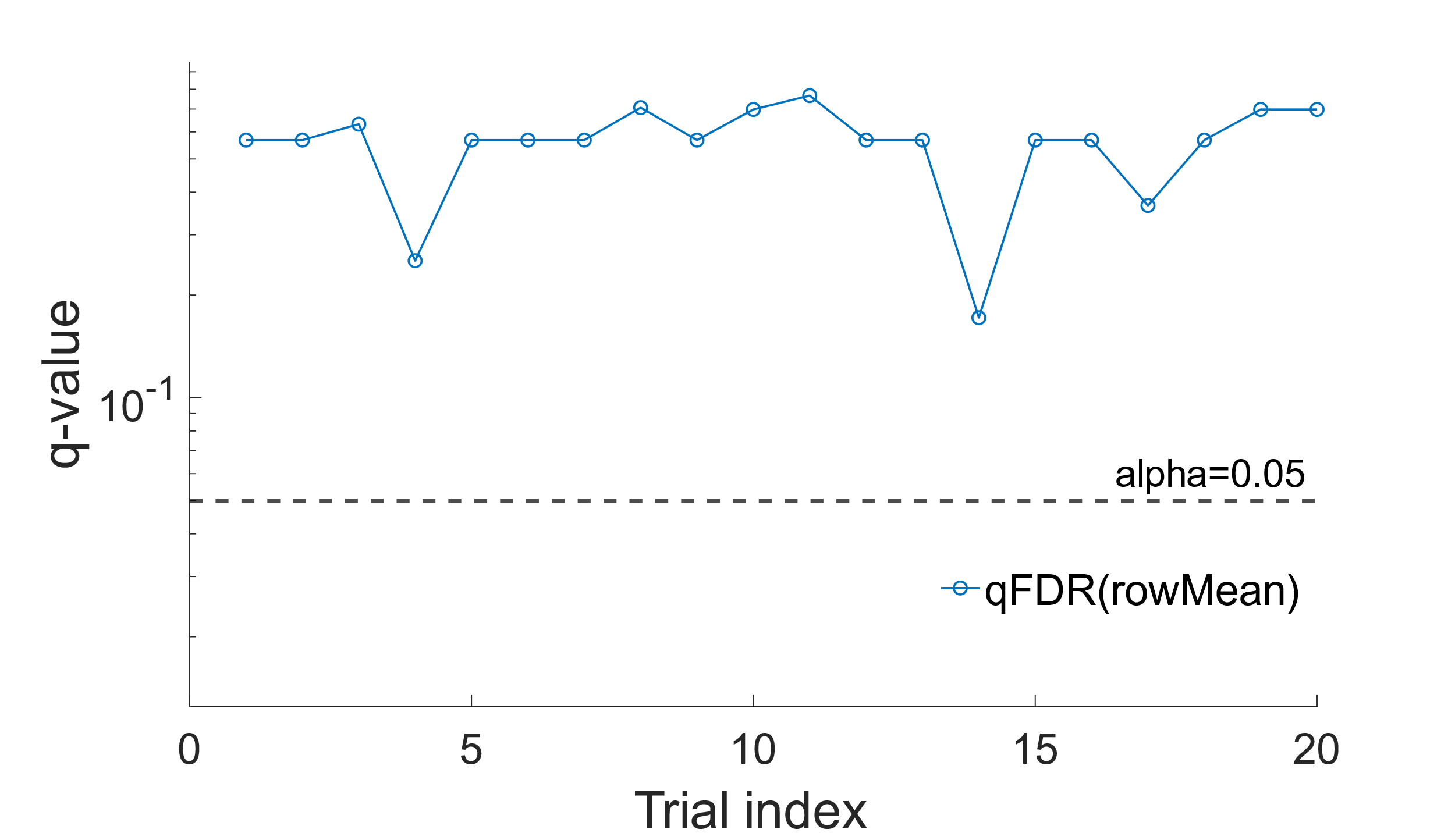

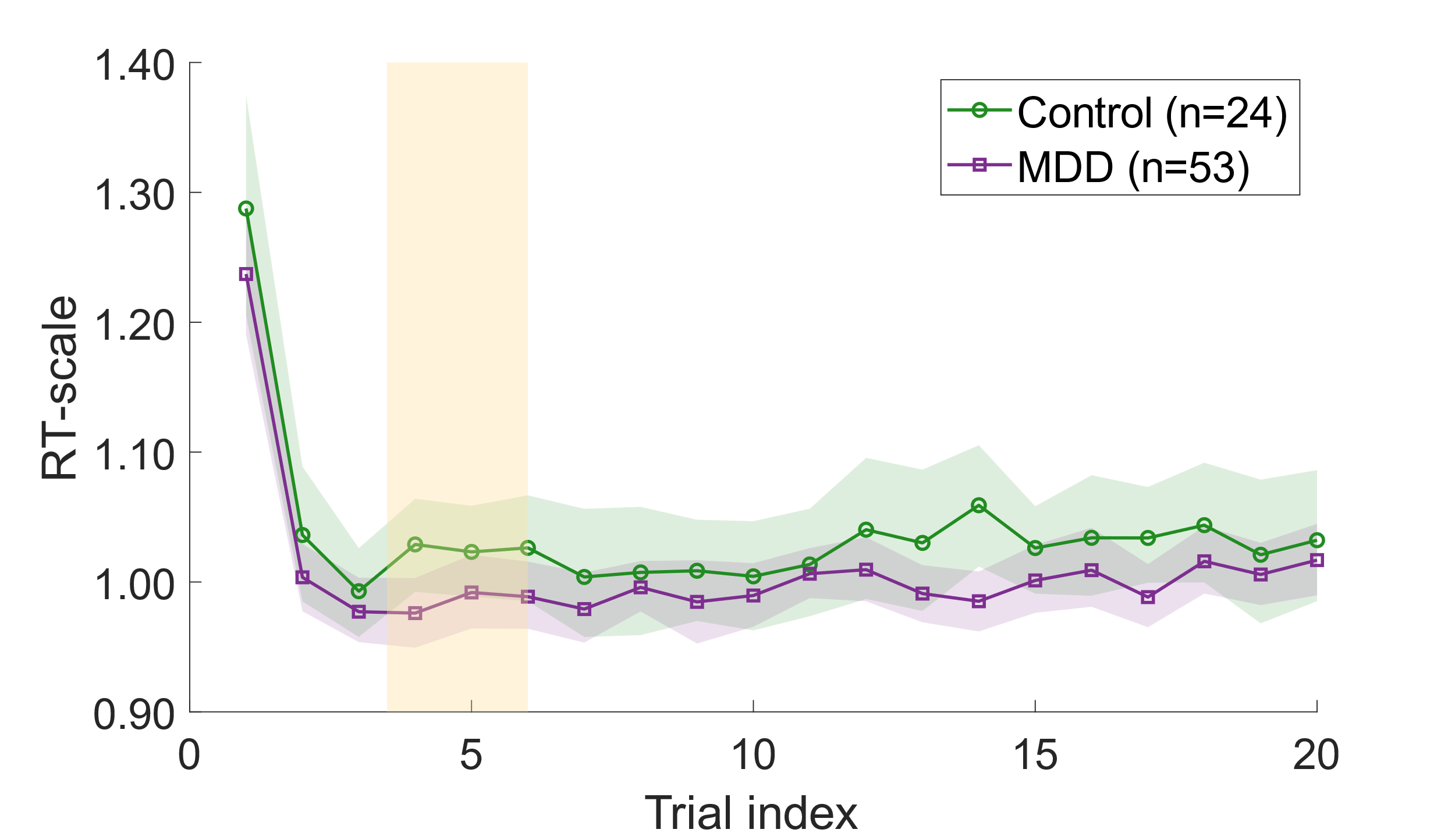

**Section 19. Trial-wise Statistical Comparison of RT-scale and PCA-derived Temporal Features.** The analyses presented in the main text suggested that SI-related temporal encoding was concentrated within a restricted subset of trials immediately following block transitions, particularly within the “Death + Me” condition. To characterize this structure more directly, we performed trial-wise statistical comparisons between SI+ and SI− participants using both the raw RT-scale trajectories and the low-dimensional latent representations derived from the nonlinear PCA framework described in the main text.

At each trial index, we compared RT-scale values between SI+ and SI− participants using two-sample t-tests with Benjamini–Hochberg false discovery rate (FDR) correction. **Fig. S8A-B** show the resulting adjusted q-values for the “Death + Me” and “Life + Me” conditions, respectively. Consistent with the temporal trajectories presented in the main text, the strongest evidence for group separation emerged during early post-switch trials within the “Death + Me” blocks, particularly around trials 4–6, although no individual trial reached significance after multiple-comparison correction. We next applied the same analysis to the PC1 trajectories derived from the nonlinear RT transformation and participant-wise uncentered PCA. In contrast to the raw RT-scale representation, the PCA-derived latent representation exhibited a sharper concentration of group-related structure at early trials, with the smallest adjusted q-value occurring at trial 4 (**Fig. S8C-D**), consistent with the temporal divergence identified throughout the main analyses.

Together, these findings indicate that SI-related information is encoded in distributed early-trial temporal dynamics rather than isolated trial-specific effects alone. Moreover, the latent PCA-based representation appears to selectively enhance the dominant temporal structure associated with SI state while suppressing unrelated single-trial variability, yielding a more concentrated representation of the underlying behavioral dynamics.

D

**Figure S8. Trial-wise Statistical Comparison Between SI+ versus SI**− **Groups.** At each trial position (1–20), we performed two-sample tests with unequal variance and report the Benjamini–Hochberg false discovery rate (FDR)–adjusted q-values. Panels **(A)** and **(B)** correspond to comparisons of the mean RT-scale in the “Death + Me” and “Life + Me” conditions, respectively. Panels **(C)** and **(D)** show the same analysis applied to the trial-wise entries of the first principal component (PC1) derived from the nonlinear transformation and uncentered PCA of the block trajectories. A horizontal dashed line indicates the nominal significance level (α = 0.05).

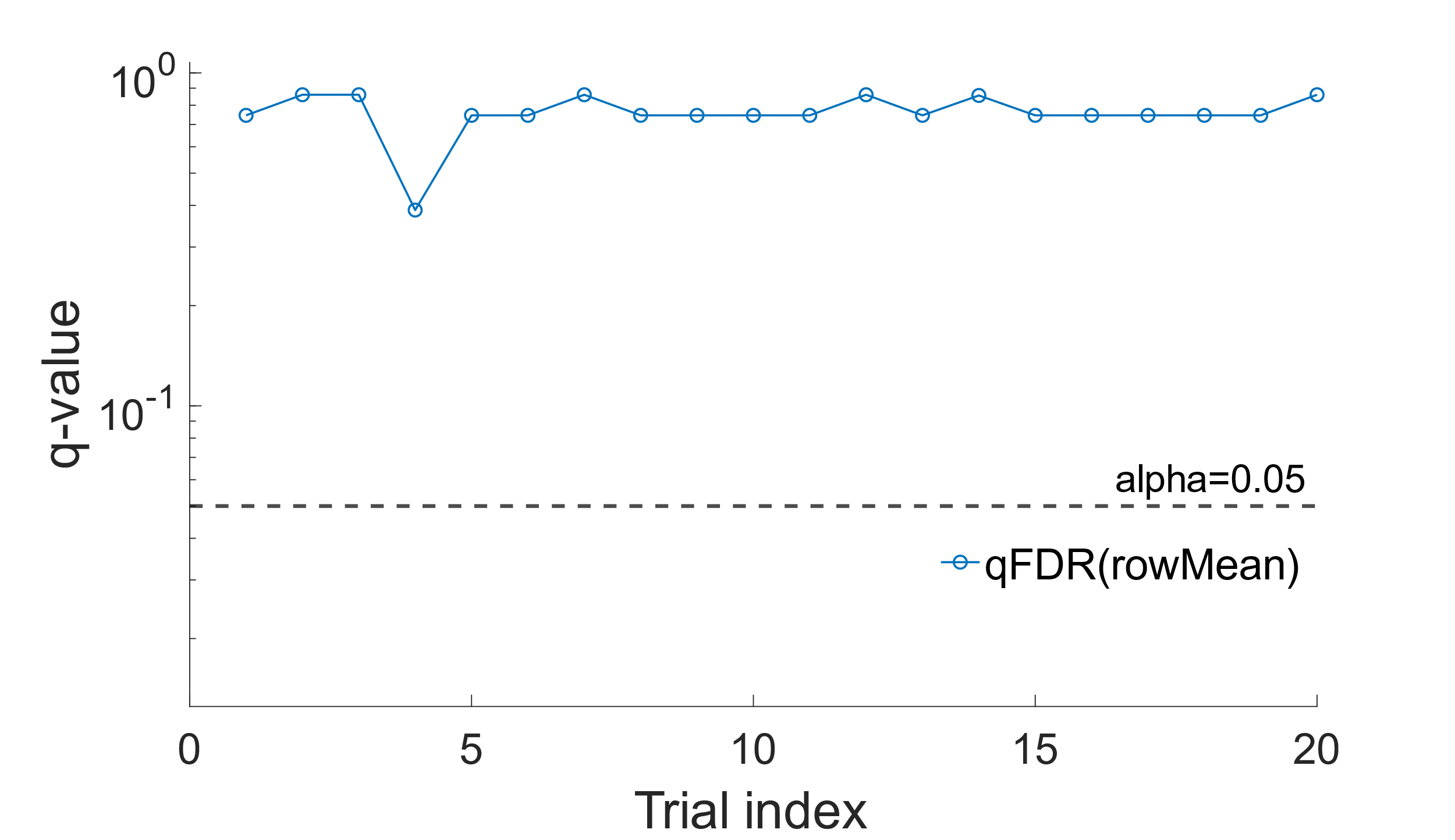

A

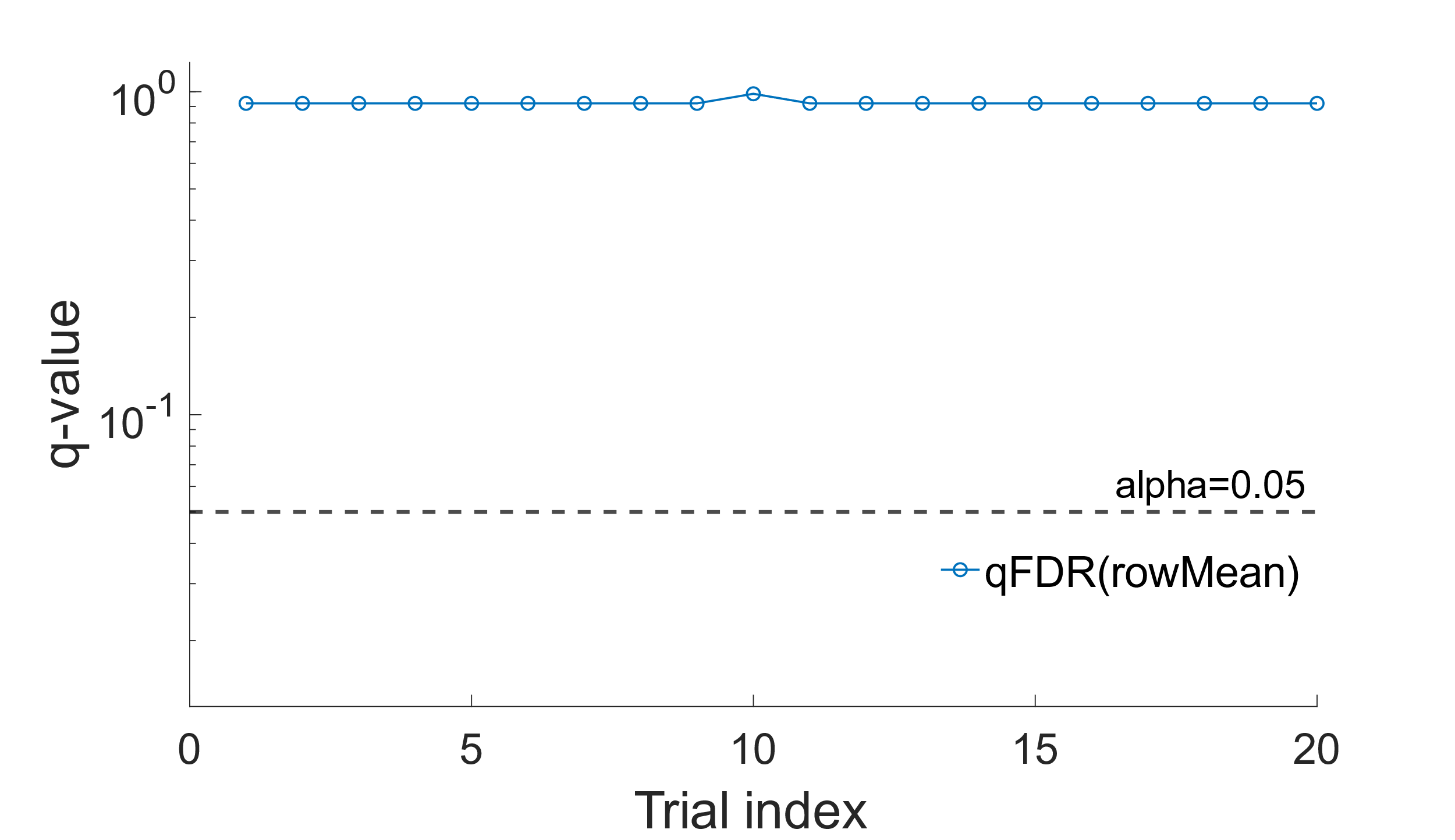

B

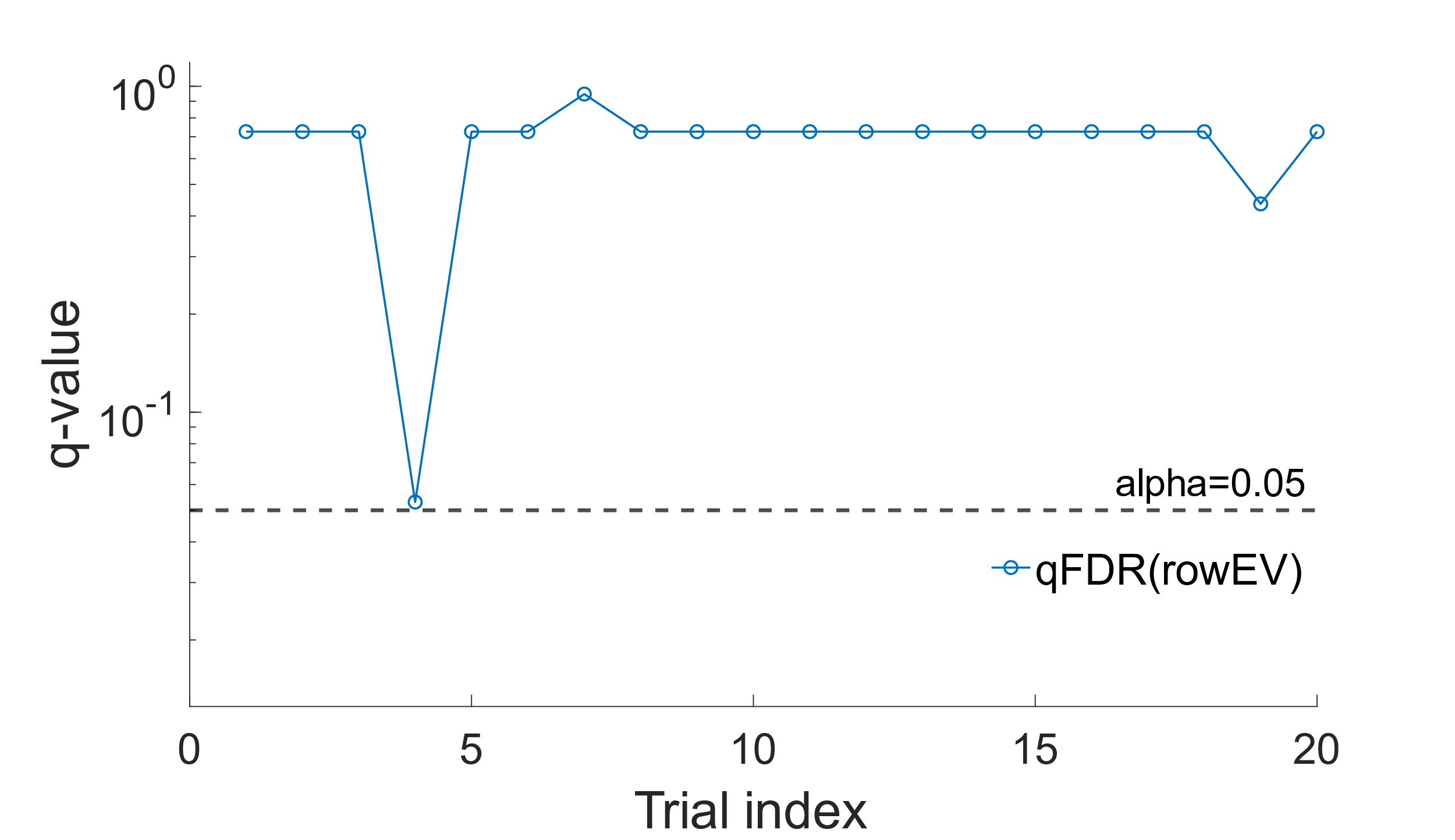

C

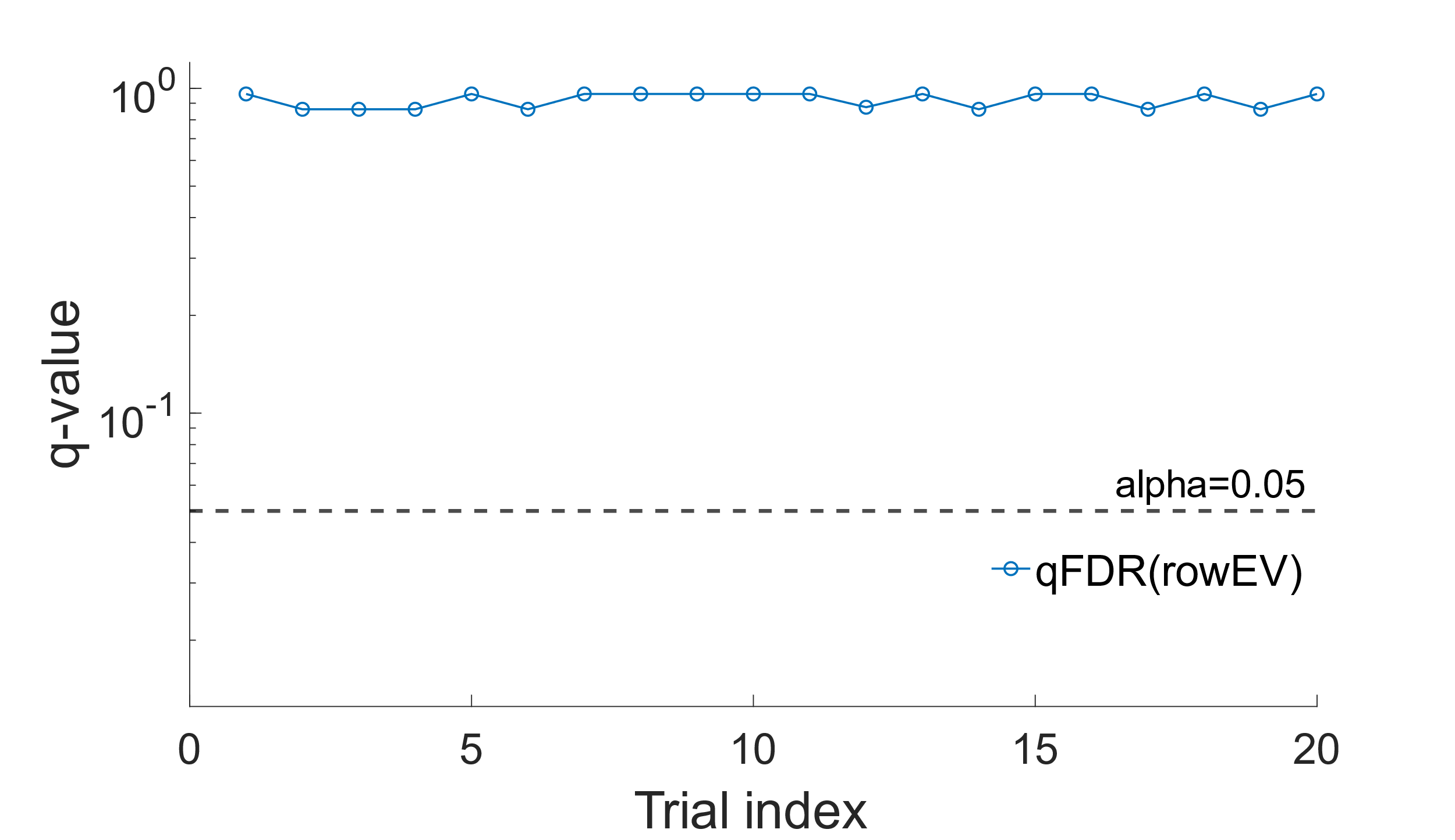
